# Prospectively validated augmented intelligence for disease-agnostic predictions of clinical success for novel therapeutics

**DOI:** 10.1101/2022.03.14.22272372

**Authors:** Bragi Lovetrue, Idonae Lovetrue

**Affiliations:** Demiurge Technologies AG, Baarerstrasse 14 6300 Zug Switzerland

## Abstract

Standalone artificial intelligence has not alleviated the long-term bottleneck of linearly extracting new knowledge from exponentially growing biological data, which has severely limited clinical success rates for drug discovery. We developed a ‘virtual patient’ augmented intelligence model that functionally reconstructed human physiology and human pathogenesis for high-fidelity simulations of drug-body interactions. We examined the clinical utility of ‘virtual patient’ in prospective predictions of clinical efficacy and safety of novel therapeutics regardless of prior clinical data availability, through a 24- month, public, prospective, large-scale, unbiased, and real-world validation study. ‘Virtual patient’ achieved 90.1% sensitivity and 82.0% precision with a 99% confidence across all major therapeutic areas, representing its capability of discovering 90.1% of all possible drug-indication pairs that could bring clinical benefits to patients, and its potential of increasing tenfold the baseline clinical success rate from 7.9% to 82.0%. ‘Virtual patient’ represents a methodological shift of drug discovery especially for age-related diseases by doing away with animal experiments whose data are hard to reproduce, virtualizing human trials whose outcomes are doomed to failure, initiating human trials whose participants are likely to benefit, and reducing R&D cycles and costs while increasing clinical efficacy and safety.

**One-Sentence Summary:** A prospectively validated ‘virtual patient’ achieved a 10.4-fold improvement in the clinical success rate for new drugs across all major diseases with 99% confidence.

---

Despite recent successes (*1, 2*), standalone artificial intelligence (AI) has not enabled life science to overcome the bottleneck of linearly extracting new knowledge from exponentially growing new data (*3*). This long-term shortage of knowledge in life science has accelerated the decline of productivity in drug discovery, even after the widespread adoption of AI for biomedical data analysis (*4*). For example, recent studies have shown that only 20% of biomedical research generated reproducible data (*5*) and fewer than 10% of animal studies successfully predicted the clinical efficacy of new drugs (*6*). It has been proposed that increasing the scale of data collection or the efficiency of knowledge extraction could address these issues (*3*).

High dimensionality and low quality are two defining characteristics of biomedical data, as biological systems involve large quantities of entities across multiple scales. These attributes make it more difficult to reproduce and structure biomedical data. Deep neural networks (DNNs) are well suited for extracting knowledge from the high-dimensional, high-quality data spaces (*2*). In contrast, biological neural networks (BNNs) are adept at extracting knowledge from low-dimensional, low-quality data spaces. As a result, neither DNNs nor BNNs alone are suitable for improving the efficiency of knowledge extraction from biological data.

The multi-faceted correspondence between DNNs and BNNs (*7*) suggests the novel possibility of combining their complementary advantages. As such, we propose adapting best practices for DNN training to a BNN, augmenting the resulting network for learning from high-dimensional data. In principle, an augmented BNN (ABNN) is well suited for learning knowledge from the high-dimensional, low-quality biological data (Fig. 1A). Despite the potential translatability of ABNNs into black-box DNNs, an ABNN is a quasi-white-box model necessarily hosted in a human brain. Hence, an ABNN is a form of human-centric augmented intelligence that could reciprocally inspire the design of improved data labels for training better-performing DNNs.

**Fig. 1.**
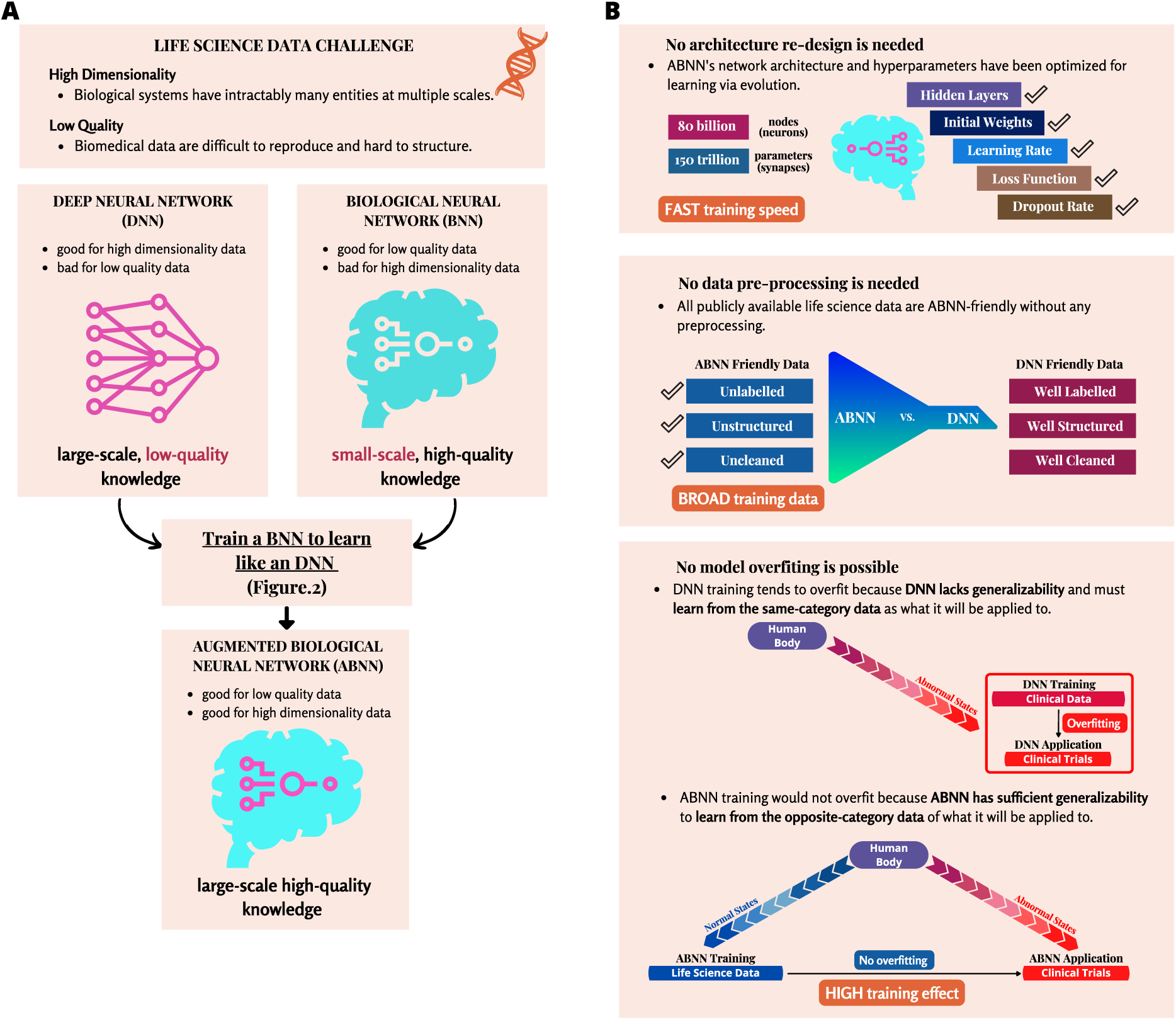
The challenge and solution of extracting knowledge from life science data. (**A**) Combining the complementary advantages of DNN and BNN: High dimensionality and low quality are two defining characteristics of life science data. Deep neural networks (DNNs) are well suited for learning knowledge from the high-dimensional, high-quality data spaces, whereas biological neural networks (BNNs) are good at learning knowledge from the low-dimensional, low-quality data spaces. The proposed augmented biological neural network (ABNN) combines the complementary advantages of DNNs and BNNs for knowledge acquisition from high-dimensional, low-quality life science data. (**B**) The training advantages of ABNNs compared with DNNs: ABNN training is far more efficient than DNN training because no time or cost is required for designing the network architecture, tuning network hyperparameters, or cleaning the training data. Furthermore, the application of an ABNN to clinical trials is also more effective than a DNN because it offers sufficient generalizability for learning from large-scale life science data, while DNNs suffer from limited generalizability for small-scale clinical data and are prone to overfitting.

There are several key advantages in training an ABNN, compared with a DNN (Fig. 1B). First, there is no need to redesign the network architecture or to tune network hyperparameters in the ABNN, since the human brain has been optimized to learn via evolution. Second, there is no need to pre-process data when training an ABNN because all publicly available life science data are interpretable by humans and thus compatible with the ABNN. This is possible even if most of the data are unlabeled, unstructured, or uncleaned, which would be inscrutable for a DNN without adequate preprocessing. Third, and more importantly, the ABNN may be immune to overfitting because it is sufficiently generalizable for learning from data that are categorically heterogeneous for its intended application. In contrast, overfitting is somewhat inevitable with a DNN because it lacks this generalizability and cannot learn from data that are not perfectly homogeneous for a given application (Fig. 1B). In the context of life science and drug discovery, an ABNN could be trained with biological data concerning normal physiological states, and then tested solely with clinical data regarding abnormal pathological states. The resulting ABNN could directly extrapolate from the normal states of human physiology to the abnormal states of human diseases, even before research data become available for new diseases like COVID-19 (*8*).

Here we introduce the first ABNN developed by training a BNN to learn like a DNN over the course of eight years (Fig. S1, A and B). Since larger DNNs typically perform better (*9, 10*), we propose that the largest ABNN could only be realized by limiting the research team size to a single brain to maximally increase the efficiency of knowledge extraction (Fig. S1, C and D; Methods section ‘Model training’). As such, best practices for DNN training were adapted to a single ABNN that learns from all-encompassing life science data in an uninterrupted pass without imposing intermediate milestones (Fig. S1E). The trained ABNN achieved exponential knowledge extraction from an exhaustive body of publicly accessible life science publications and databases, thereby constructing a foundation model (*11*) of human physiology and over 130 specific models of human diseases.

The quality of knowledge extracted with the ABNN was assessed by measuring resulting increases in the productivity of drug discovery. Inspired by the CASP challenge (*12*), which served as a benchmark for evaluating DNN performance against expert performance in protein structure prediction, we designed a real-world validation test—the <u>p</u>ublic prospective predicti<u>o</u>n of pivotal <u>o</u>ngoing <u>c</u>linical trial success <u>o</u>utcomes at large <u>s</u>cale (PROTOCOLS) test. This test provided a benchmark for comparing ABNN and expert performance in clinical success prediction (Fig. 2, A to C; Methods section ‘Validation design’).

**Fig. 2.**
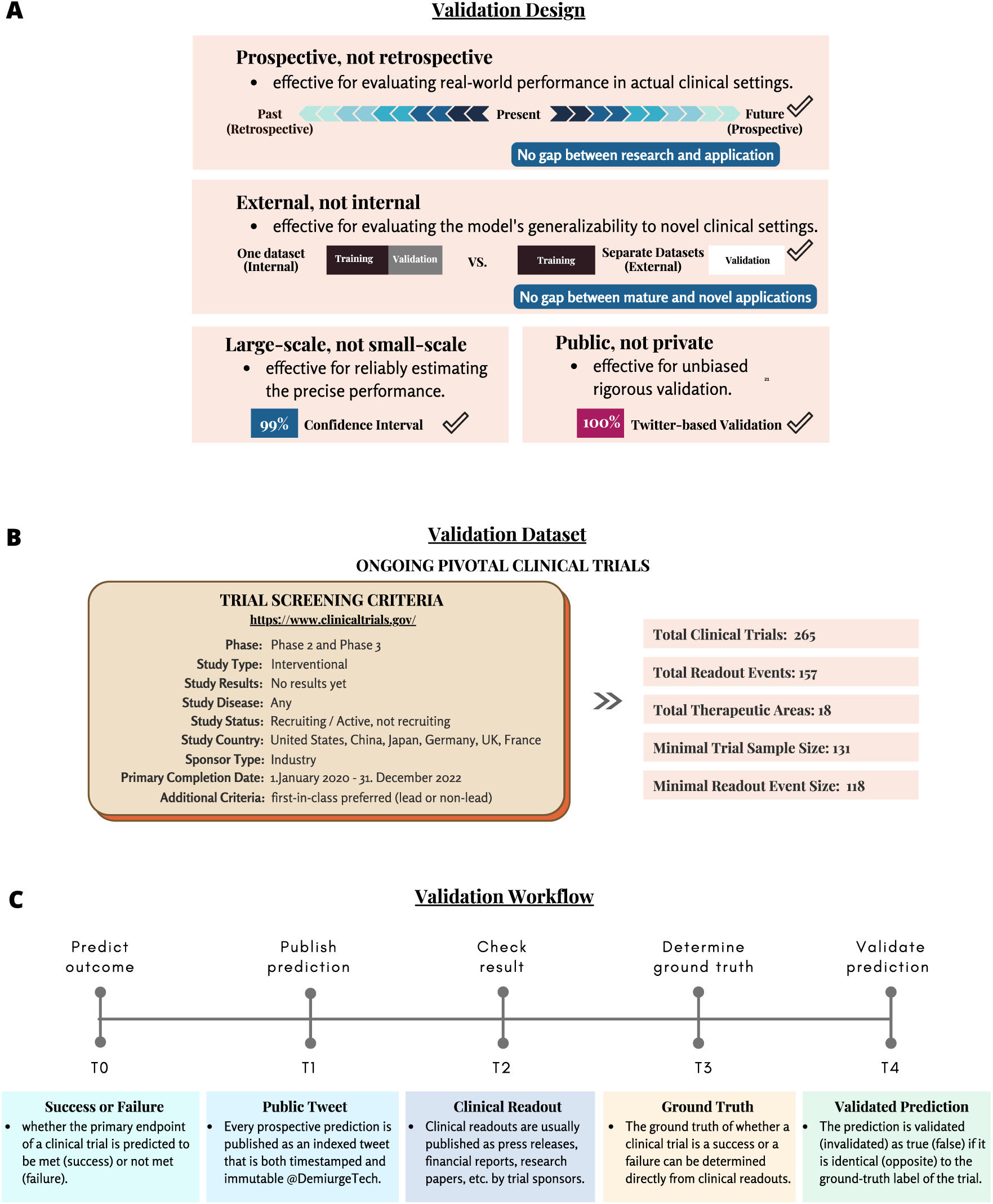
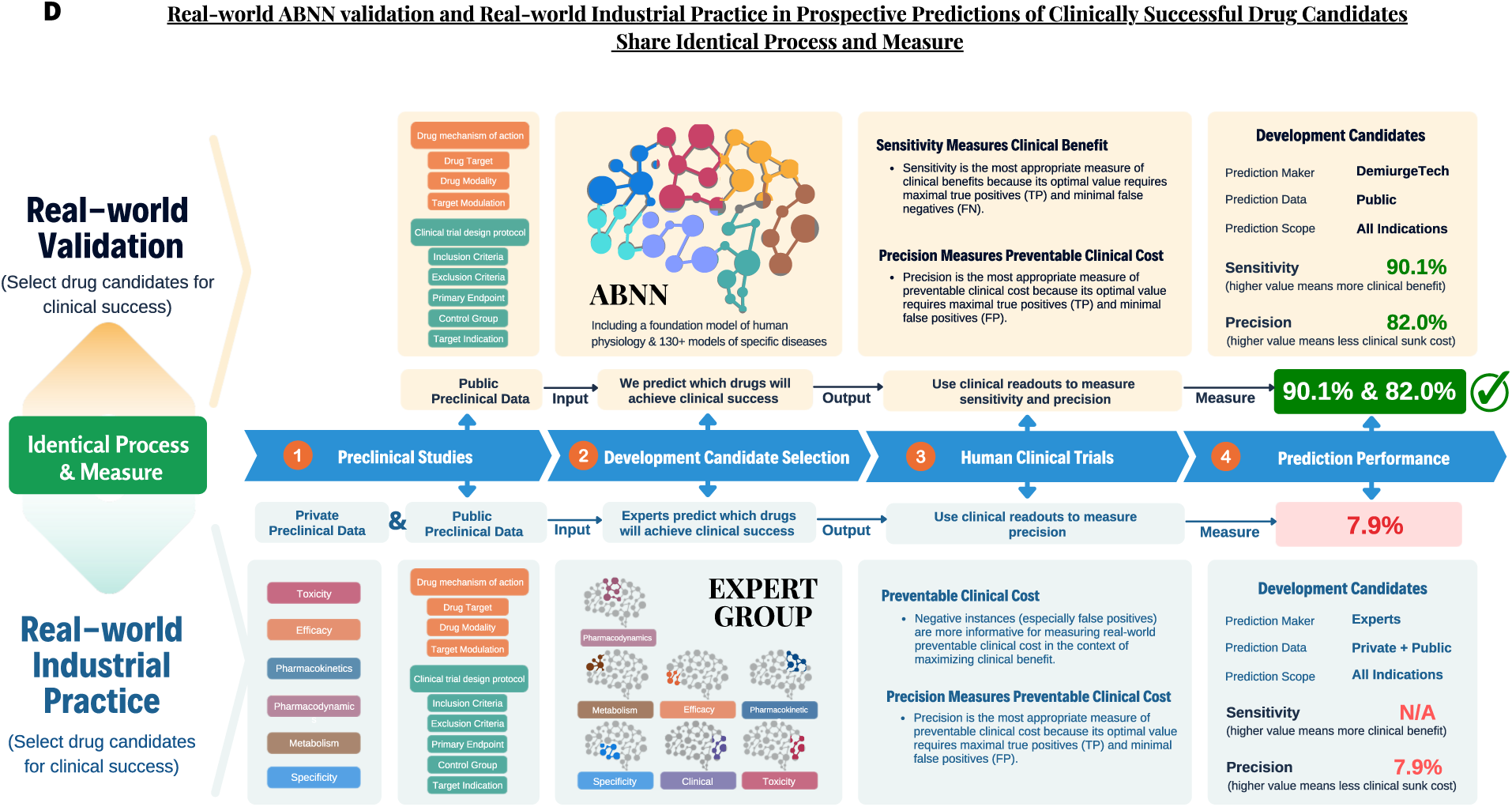
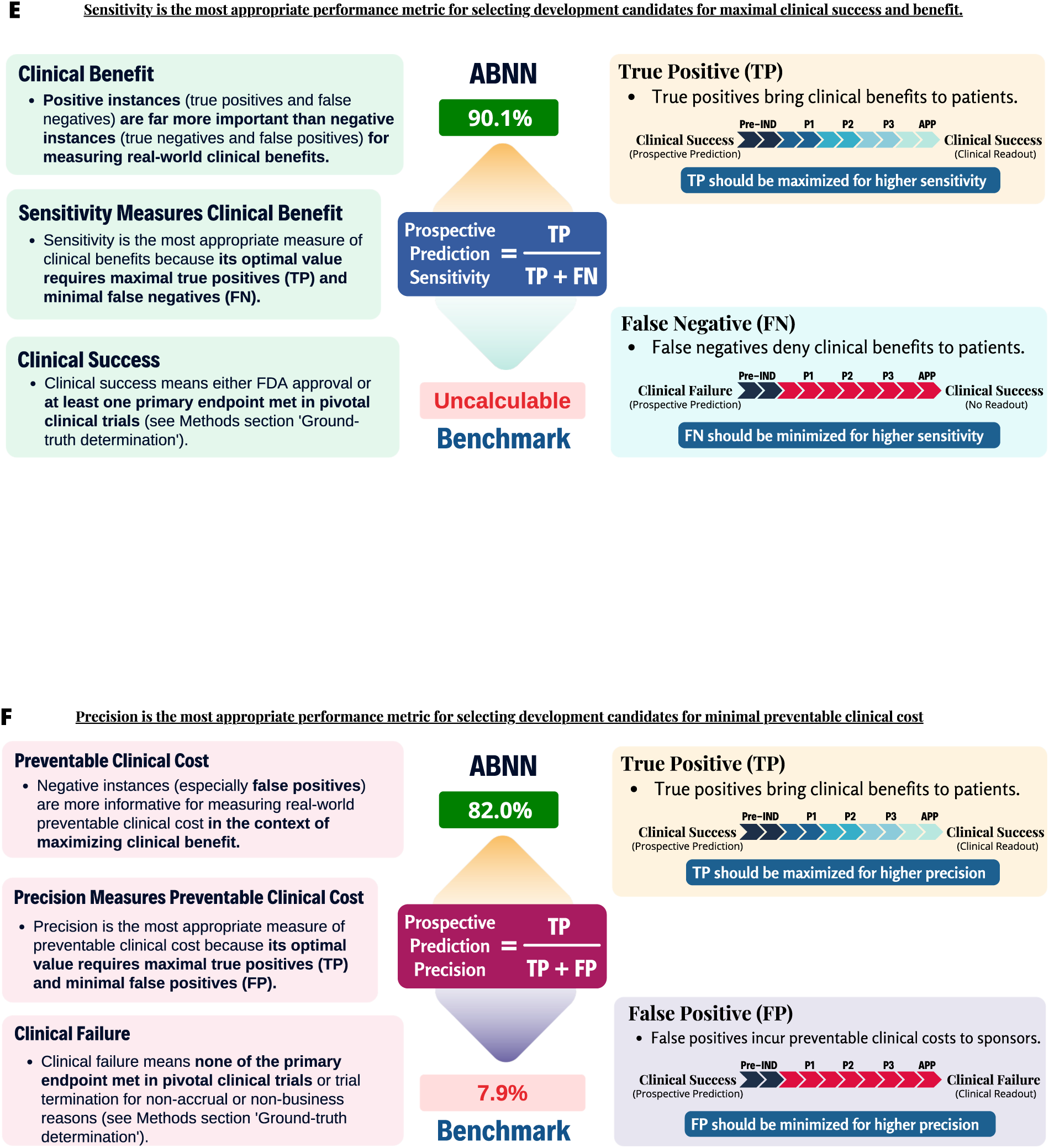
A real-world, large-scale, public, prospective, external validation of the trained ABNN. (**A**) Prospective validation is far more rigorous than retrospective validation for examining ABNN performance in real-world clinical settings. Large-scale validation is also more rigorous than small-scale validation for enabling an unbiased estimation of the actual predictive power of the ABNN. External validation using separate datasets (independent of the training datasets) is more rigorous than internal validation using held-out portions of the training data, in evaluating the generalizability of the ABNN for targeted clinical applications. Public validation is more rigorous than private validation for ruling out all the possibility of biasing performance estimates. (**B**) The list of screening criteria used to identify pivotal clinical trials for ABNN validation. ABNN performance was evaluated using the public prospective predictions of pivotal ongoing clinical trial success outcomes at large scale (PROTOCOLS), the most rigorous external validation test to date of a clinical prediction model with a binary outcome. A total of 265 prospective predictions were published to satisfy the minimum sample size requirement of 131 and evaluate prospective prediction sensitivity, prospective prediction precision, F1 score, accuracy, and specificity at 99% confidence. Similarly, 157 readouts were accumulated to meet the minimum event size requirement of 118 (see Methods section ’Statistical analysis’). (**C**) The workflow for every validation step from beginning to end (see Methods section for details). Twitter was used to meet the stringent criteria in PROTOCOLS, providing immutable, timestamped, and indexed tweets to construct a publicly verifiable track record on the publicly accessible account of @DemiurgeTech. The ABNN was used to made prospective predictions of pivotal clinical trial outcomes for all human diseases, excluding rare neurodevelopmental, rare skeletomuscular, and hematological disorders. There was no selection of lead over non-lead indications, though non-lead indications have far lower success rates. There are a few hundred new pivotal clinical trials whose protocols are publicly accessible in a centralized database, as required by FDA, and whose clinical readouts are typically published by sponsors in a standardized format. We calculated prospective prediction precision with a 99% confidence interval using a ground truth determined by actual clinical readouts. (**D**) Real-world validation is critical to the objective assessment of ABNN’s potential for improving the efficiency of global drug research and development. As the foundation model in the ABNN is intended to functionally reconstruct nearly all major diseases that could occur to a single human patient, clinical success prediction is the most effective validation method for the ABNN. The clinical efficacy and safety of novel drugs were evaluated using mechanisms of action, to prevent drug developers from initiating pivotal clinical trials that are doomed to failure. Several key go/no-go decision steps are involved in drug development, as illustrated in Steps 1-4 in the figure. In Step 1, preclinical studies aim to determine which drug candidates are developable and generate sufficient data indicative of efficiency and safety in humans for each developable candidate. Biopharmaceutical companies have a considerable data advantage over the ABNN in this step as they have exclusive access to internally generated private data in addition to public data. In contrast, the ABNN only has access to public data of drug mechanisms of action (i.e., drug targets, drug modality, and target modulation) and clinical trial design protocols (i.e., inclusion criteria, exclusion criteria, primary endpoint, control groups, and target indication). Step 2 is a critical go/no-go decision step in drug development because only a subset of developable drug candidates (with highest clinical success potential) can be selected to transition from non-clinical to clinical development, due to the prohibitive costs of human clinical trials. Best practices in the biopharmaceutical industry involve a group of committed experts using private and public preclinical data to predict which drug candidates will achieve clinical success. In Step 3, clinical trials are then initiated only for those drug candidates that are prospectively predicted to be clinically successful. In Step 4, in the current practices of the biopharmaceutical industry, the resulting prospective prediction precision is only 7.9% (i.e., for every 100 clinical programs that experts prospectively predict to achieve clinical success, only 7.9 are actual successful) and the resulting prospective prediction sensitivity is incalculable because false negatives would not be greenlit to initiate clinical trials in the first place. From Step 2 to 4, the real-world PROTOCOLS validation test was designed to mirror real-world industrial practices, to ensure the transferability of ABNN performance from validation to application. The ABNN solely uses public preclinical data to predict which drug candidates will achieve clinical success (Step 2). Clinical readouts were tracked and recorded for those drug candidates that are prospectively predicted to be clinically successful (Step 3). The resulting prospective prediction sensitivity is 90.1% (i.e., for every 100 clinical programs that achieved actual clinical success, 90.1 were correctly predicted by the ABNN). The resulting prospective prediction precision was 82.0% (i.e., for every 100 clinical programs that were prospectively predicted to achieve clinical success by the ABNN, 82 were actually successful) (Step 4). *These indications cover all diseases except for hematological disorders, rare neurodevelopmental disorders, and rare musculoskeletal disorders. (**E**) Sensitivity is the most appropriate performance metric for selecting development candidates for maximal clinical success and benefit. Positive instances (true positives and false negatives) are far more important than negative instances (true negatives and false positives) for measuring real-world clinical benefits. Sensitivity is the most appropriate measure of clinical benefits because its optimal value requires maximal true positives (TP) and minimal false negatives (FN). Clinical success means either FDA approval or at least one primary endpoint met in pivotal clinical trials (see Methods section ’Ground-truth determination’). (**F**) Precision is the most appropriate performance metric for selecting development candidates for minimal preventable clinical cost. Precision is the most appropriate measure of preventable clinical cost because its optimal value requires maximal true positives (TP) and minimal false positives (FP). Clinical failure means none of the primary endpoint met in pivotal clinical trials or trial termination for non-accrual or non-business reasons (see Methods section ’Ground-truth determination’).

The rationale for the PROTOCOLS test is that pivotal clinical trials (phase 2 and phase 3) are statistically powered to assess a drug’s clinical success by evaluating its clinical efficacy and safety, which is largely determined by the underlying biological processes that can be distilled into scientific knowledge. Historical clinical success rates for investigational drugs are well-documented (*13, 14*) and provide an accurate baseline for evaluating ABNN performance in the PROTOCOLS test.

The PROTOCOLS validation test was used to evaluate the real-world performance of the ABNN in actual clinical settings with an emphasis on generalizability to novel drug-disease pairs, even with scarce or uninformative prior clinical data (Fig. 2, A and B). As such, there is little difference between the validation scenario of the ABNN in the PROTOCOLS validation test and the application scenario of the ABNN in real-world workflow of drug discovery and development (Fig. 2D) (*15, 16*).

Clinical success prediction is a critical go/no-go decision step in drug development because only a subset of developable drug candidates, with the highest clinical success potential, are selected to transition from non-clinical to clinical development, due to the prohibitive cost of human clinical trials (*17*). As a best practice in the biopharmaceutical industry (*18*), a group of committed experts review both publicly available and privately accessible preclinical data to predict which drug candidates could achieve clinical success. Human clinical trials are then initiated only for those drug candidates that are prospectively predicted to be clinically successful (Fig. 2D).

It should be noted that positive instances (true positives and false negatives) are far more important than negative instances (true negatives and false positives) for measuring real-world clinical benefits of novel therapeutics (Fig. 2E). True positives are also more interesting than true negatives because the historical clinical success rate for investigational drugs entering human studies is only 7.9% across all diseases (*13*). As such, false negatives have a substantial clinical impact because they not only deny clinical benefits to patients, but also eliminate sizable revenue opportunities. Prospective prediction sensitivity is the most appropriate measure of clinical benefits because its optimal value requires maximal true positives and minimal false negatives (Fig. 2E; Methods section ‘Metrics definition’).

On the other hand, negative instances (especially false positives) are more informative for measuring real-world preventable clinical cost in the context of maximizing clinical benefit (Fig. 2F). As such, false positives have only a marginal impact on clinical benefit because human clinical trials can still fail without doing much harm to patients, yet they have a significant impact on preventable clinical cost because drug developers could have refrained from initiating pivotal clinical trials that are doomed to failure. Prospective prediction precision is the most appropriate measure of preventable clinical cost because its optimal value requires maximal true positives and minimal false positives (Fig. 2F; Methods section ‘Metrics definition’).

Every investigational new drug greenlit to enter first-in-human studies is then genuinely believed by committed experts to be capable of achieving clinical success. As such, the clinical success rate from phase 1 to approval (*14*) captures false positives and is construed as the prospective prediction precision by experts to benchmark the prospective prediction precision by the ABNN in PROTOCOLS validation test (Fig. 2, D and F; Methods section ‘Metrics definition’). In contrast, as none of the drug candidates ruled out by experts would ever be greenlit to enter first in human studies, no false negatives could be discovered for experts from historical clinical trial data. The prospective prediction sensitivity by experts is thus incalculable in current practices of drug development for benchmarking the prospective prediction sensitivity by the ABNN (Fig. 2, D and E; Methods section ‘Metrics definition’).

Accordingly, we chose to report prospective prediction sensitivity (recall) in the context of F1 score, and prospective prediction precision against the expert benchmark as the most appropriate performance metrics for PROTOCOLS validation test (*19*) to reflect the descending priorities (maximize true positives > minimize false negatives > minimize false positives > maximize true negatives) in actual clinical settings to deliver maximal clinical benefit to patients at minimal clinical cost to sponsors (Methods section ‘Metrics definition’ and ‘Model training’) (*20, 21*). We also report other standard measures (accuracy, specificity, etc.) to provide a balanced view of ABNN performance (Table 1, and Table S1).

**Table 1.** Performance metrics with statistical significance.

| **Validation Subgroups** | **N** | **Metric** | **Performance** | **Significance** | **CI (%)** | **p-value** |
| --- | --- | --- | --- | --- | --- | --- |
| All Indications | 157 | F1-Score | 0.86 | 99% | (0.79, 0.92) | 0.0001 |
| All First-in-class Indications | 126 | F1-Score | 0.85 | 99% | (0.76, 0.92) | 0.0001 |
| All Age-related Indications | 119 | F1-Score | 0.85 | 99% | (0.76, 0.92) | 0.0001 |
| All Infectious Disease Indications | 49 | F1-Score | 0.78 | 90% | (0.65, 0.86) | 0.0012 |
| All Oncology Indications | 48 | F1-Score | 0.85 | 90% | (0.77, 0.90) | 0.0004 |
| All NME Indications | 77 | F1-Score | 0.82 | 90% | (0.73, 0.88) | 0.0002 |
| All Biologics Indications | 68 | F1-Score | 0.87 | 90% | (0.80, 0.91) | 0.0002 |
| All Indications | 157 | Sensitivity | 90.1% | 99% | (80.0, 96.9) | 0.001 |
| All First-in-class Indications | 126 | Sensitivity | 92.4% | 99% | (80.8, 99.2) | 0.009 |
| All Age-related Indications | 119 | Sensitivity | 91.0% | 99% | (79.1, 98.4) | 0.005 |
| All Infectious Disease Indications | 49 | Sensitivity | 88.9% | 90% | (68.3, 95.4) | 0.050 |
| All Oncology Indications | 48 | Sensitivity | 88.9% | 90% | (75.7, 94.3) | 0.015 |
| All NME Indications | 77 | Sensitivity | 85.7% | 90% | (71.9, 92.2) | 0.009 |
| All Biologics Indications | 68 | Sensitivity | 91.5% | 90% | (80.8, 95.7) | 0.015 |
| All Indications | 157 | Precision | 82.0% | 99% | (70.8, 90.7) | 0.0001 |
| All First-in-class Indications | 126 | Precision | 78.2% | 99% | (64.8, 88.8) | 0.0001 |
| All Age-related Indications | 119 | Precision | 79.2% | 99% | (65.9, 89.7) | 0.0001 |
| All Infectious Disease Indications | 49 | Precision | 69.6% | 90% | (51.8, 81.6) | 0.0035 |
| All Oncology Indications | 48 | Precision | 82.1% | 90% | (68.9, 89.3) | 0.0030 |
| All NME Indications | 77 | Precision | 78.9% | 90% | (65.4, 87.0) | 0.0018 |
| All Biologics Indications | 68 | Precision | 82.7% | 90% | (71.6, 89.1) | 0.0011 |
| All Indications | 157 | Accuracy | 82.8% | 99% | (74.2, 89.8) | 0.0001 |
| All First-in-class Indications | 126 | Accuracy | 82.5% | 99% | (72.8, 90.3) | 0.0001 |
| All Age-related Indications | 119 | Accuracy | 81.5% | 99% | (71.3, 89.7) | 0.0001 |
| All Infectious Disease Indications | 49 | Accuracy | 81.6% | 90% | (70.1, 88.4) | 0.0020 |
| All Oncology Indications | 48 | Accuracy | 77.1% | 90% | (65.1, 84.9) | 0.0004 |
| All NME Indications | 77 | Accuracy | 83.1% | 90% | (74.4, 88.6) | 0.0002 |
| All Biologics Indications | 68 | Accuracy | 80.9% | 90% | (71.3, 87.0) | 0.0002 |
| All Indications | 157 | Specificity | 72.7% | 99% | (57.5, 85.3) | 0.0001 |
| All First-in-class Indications | 126 | Specificity | 71.7% | 99% | (55.6, 85.0) | 0.0001 |
| All Age-related Indications | 119 | Specificity | 69.2% | 99% | (51.8, 83.9) | 0.0001 |
| All Infectious Disease Indications | 49 | Specificity | 77.4% | 90% | (62.1, 86.4) | 0.0040 |
| All Oncology Indications | 48 | Specificity | 41.7% | 90% | (23.4, 64.2) | 0.0080 |
| All NME Indications | 77 | Specificity | 81.0% | 90% | (68.3, 88.3) | 0.0018 |
| All Biologics Indications | 68 | Specificity | 57.1% | 90% | (39.7, 72.3) | 0.0016 |

The PROTOCOLS validation test was designed to mirror the real-world industrial practices and minimize the performance gap between validation and application (Fig. 2D). The included ABNN accessed a publicly available subset of expert-accessible preclinical data, used to predict which drug candidates would achieve clinical success. Clinical trial readouts were tracked for all drug candidates that had been prospectively predicted to be either clinically successful or unsuccessful (Fig. 2D, and Fig. S3; Methods section ‘Prediction generation’). The real-world industrial practice has yielded a prospective prediction precision of 7.9% (Methods section ‘Baseline performance’) (*14*), which is only slightly better than chance (*22*). In contrast, the ABNN followed identical procedures with much fewer data yet yielded a prospective prediction precision of 82.0%, representing a 10.4-fold improvement over the baseline in realistic settings (Fig. 2D).

## RESULTS

The PROTOCOLS validation test comprises 265 then-ongoing real-world pivotal clinical trials following a predefined set of screening criteria (Fig. 2B and Methods section ‘Validation data collection’).

Validation clinical trials and newly initiated clinical trials since 2020 were shown to follow identical distributions across all major therapeutic areas (two-sample Kolmogorov-Smirnov test, statistic = 0.333; P = 0.73; Fig. 3A). PROTOCOLS thus provided an unbiased evaluation of disease-agnostic performance for the ABNN.

**Fig. 3.**
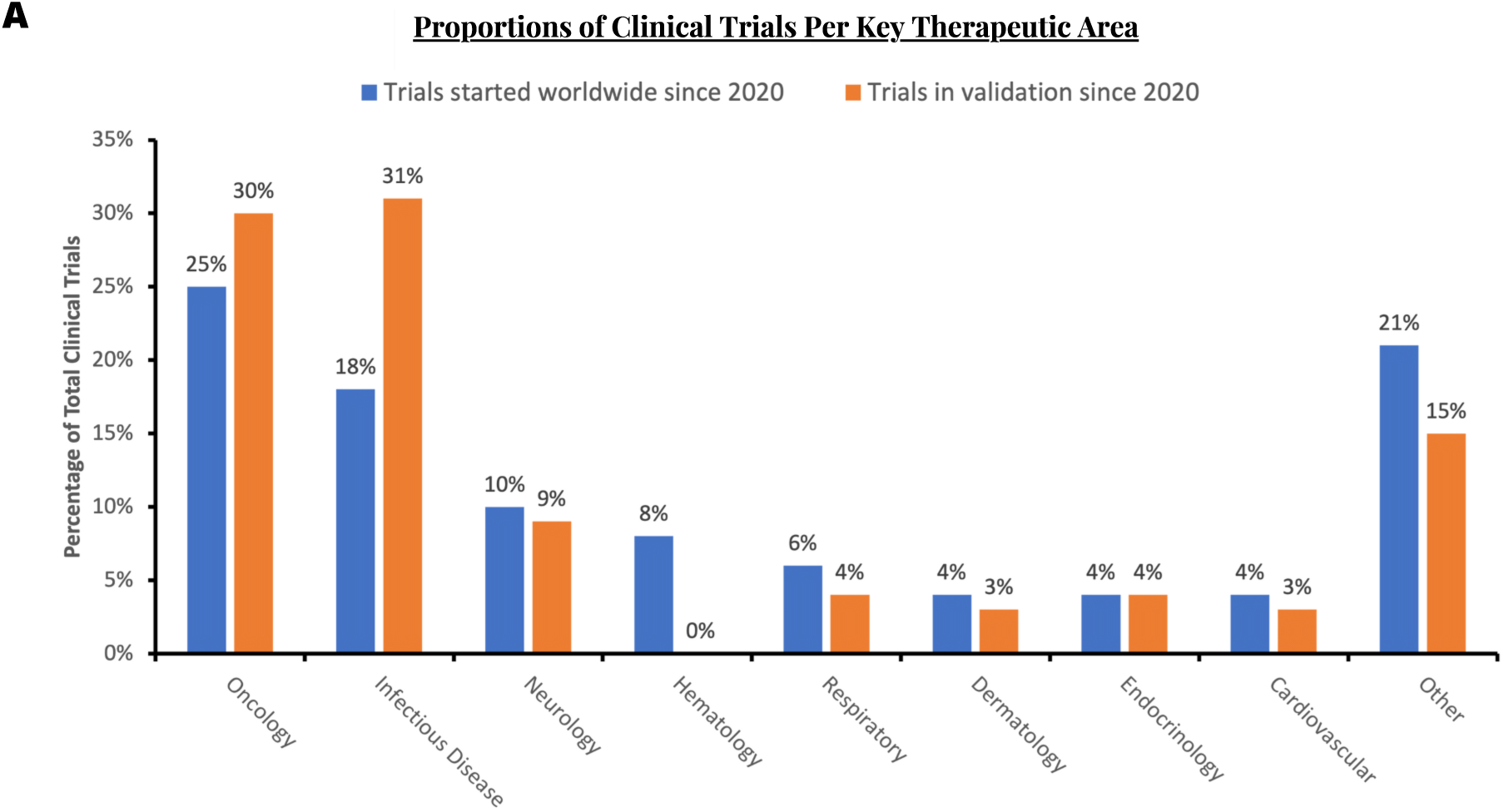
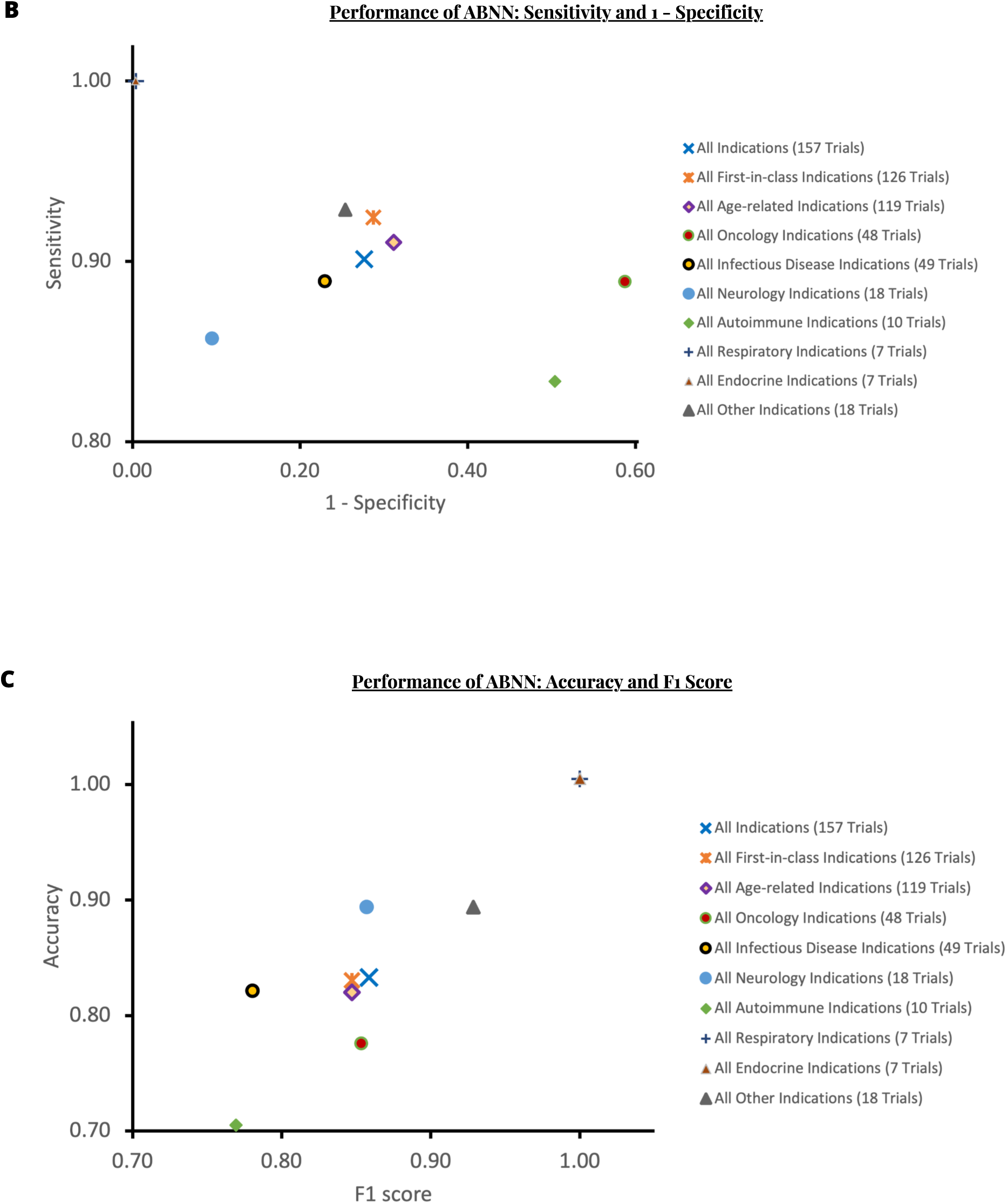
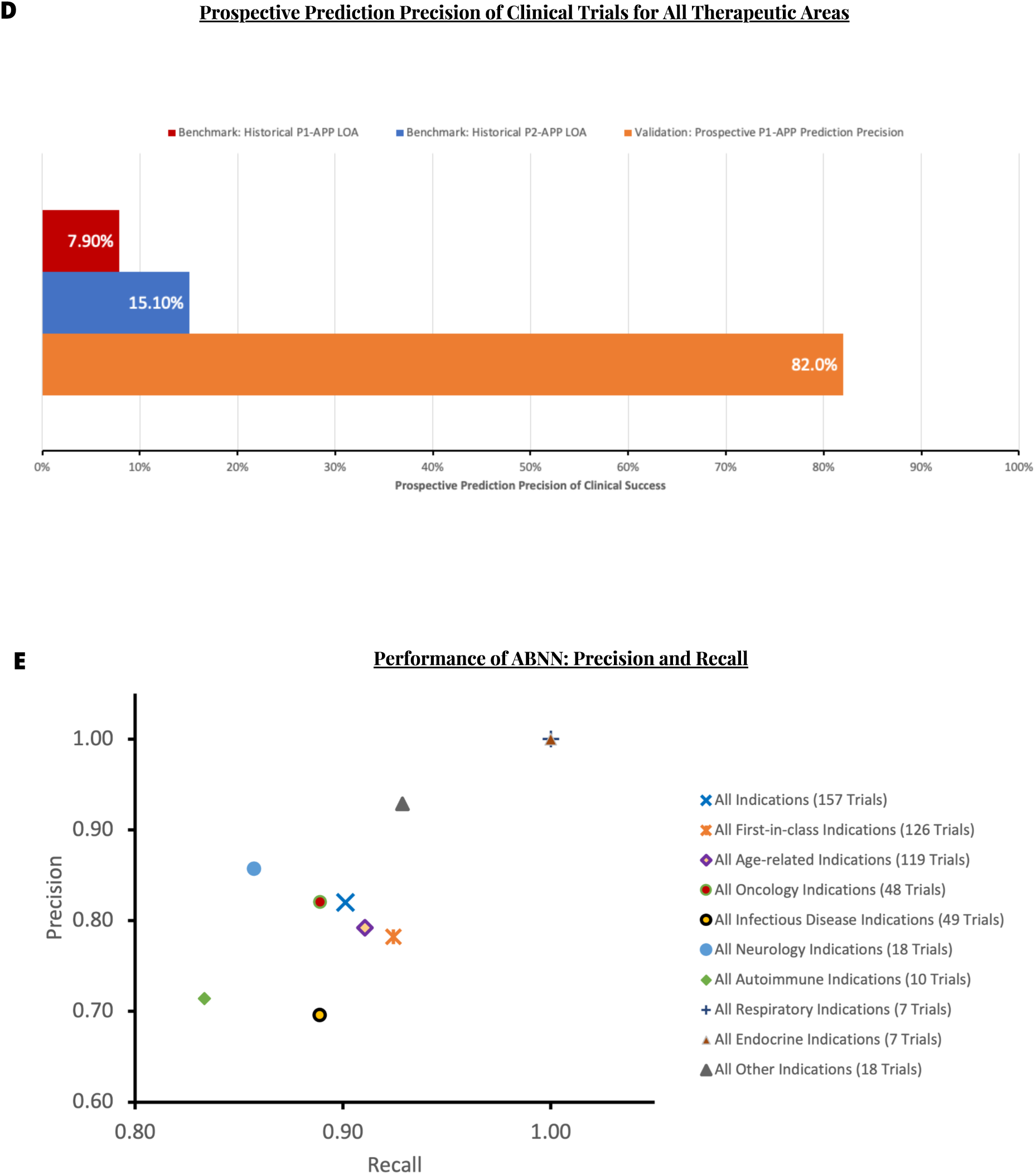
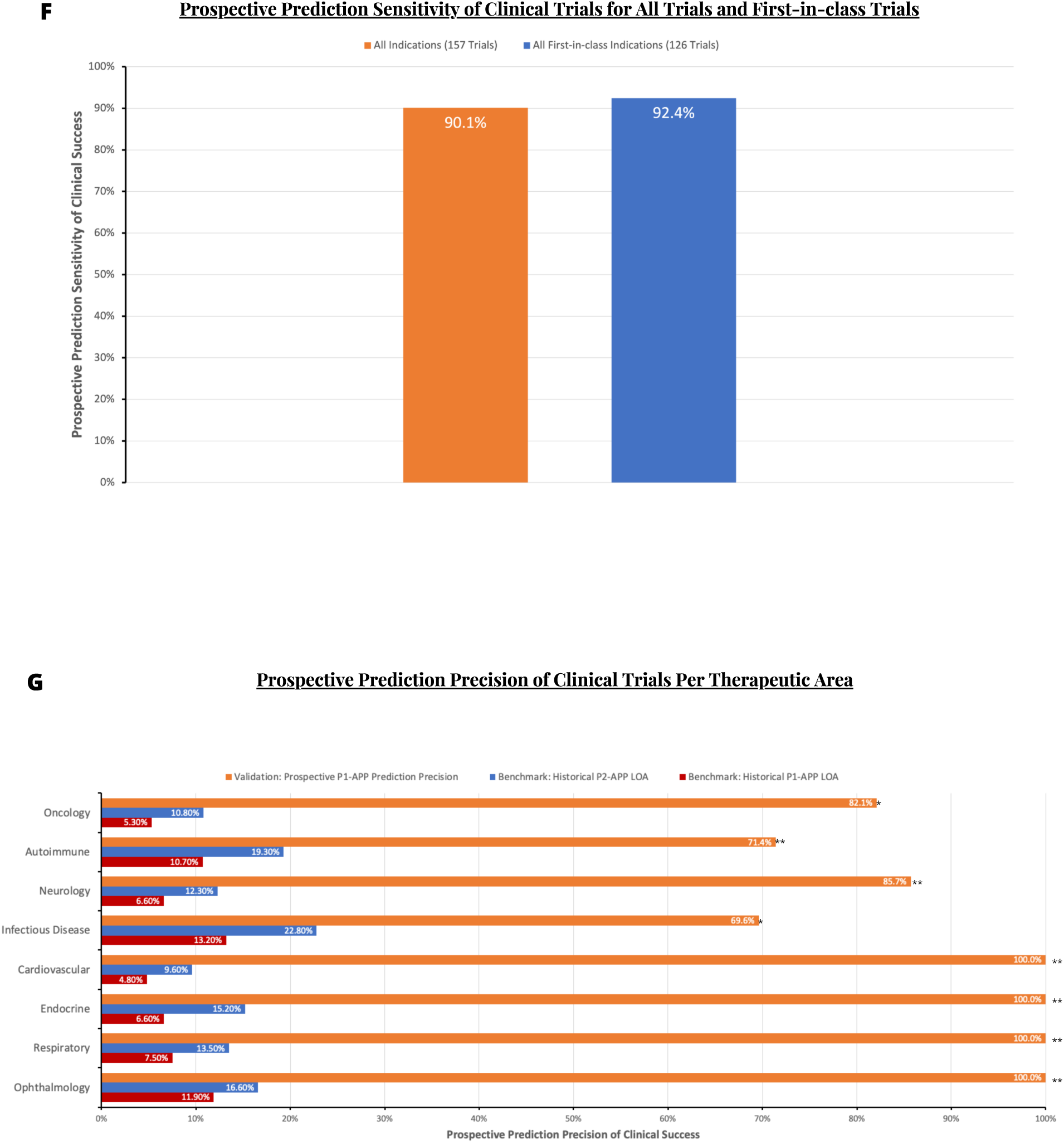
Prospective prediction performance metrics across therapeutic areas. (**A**) A two-sample Kolmogorov-Smirnov test was used as the standard method to evaluate the direct translatability between prospective validation and clinical applications. Validation clinical trials and all clinical trials initiated since 2020 followed identical distributions (statistic = 0.333 p-value = 0.73). (**B**) ABNN performance as measured by sensitivity and 1-specificity (see Table 1 for more details). (**C**) ABNN performance as measured by accuracy and F1 score (see Table 1 for more details). (**D**) PROTOCOLS test involved a realistic baseline precision of 7.90% (Historical P1-APP LOA: historical likelihood of success from phase 1 to approval) and a conservative baseline precision of 15.1% (Historical P2-APP LOA: historical likelihood of success from phase 2 to approval) to benchmark ABNN performance (see Methods section ’Baseline performance’ for details). The overall prospective prediction precision for pivotal clinical trials in the PROTOCOLS validation was 82.0% (99% CI 70.8%, 90.7%; P < 0.0001). **(E)** ABNN performance as measured by precision and recall across all validation subgroups are strongly positively correlated (exact Pearson coefficient = 0.808) (see Table 1 for more details). **(F)** Prospective prediction sensitivity for first-in-class pivotal clinical trials was superior to that of all pivotal clinical trials (Δ = 2.3%; 99% CI -0.92%, 5.52%; P < 0.002; 2% margin for non-inferiority) for all therapeutic areas, excluding rare neurodevelopmental, rare skeletomuscular, and hematological disorders (see Methods section ’Model training’ for details). (**G**) PROTOCOLS test utilized a realistic baseline precision (Historical P1-APP LOA: historical likelihood of success from phase 1 to approval) and a conservative baseline precision (Historical P2-APP LOA: historical likelihood of success from phase 2 to approval) to benchmark ABNN performance (see Methods section ’Baseline performance’ for details). Remarkably, the ABNN achieved 82.1% prospective prediction precision for oncology (90% CI 68.9%, 89.3%; P < 0.003). *The minimum sample and event size requirements for a 90% confidence have been met (see Methods section ‘Statistical analysis’). **The minimum sample and event size requirements for 90% confidence have not been met (see Methods section ‘Statistical analysis’).

Minimum sample size (131) and event size (118) requirements were determined for evaluating ABNN performance with 99% confidence (Fig. 2B and Methods section ‘Statistical analysis’) (*23*). Both requirements were met as clinical readouts were available from 157 of 265 predictions by the cutoff date (Methods section ‘Clinical readout’). A ground truth was then determined for each readout and was then applied to validation of the corresponding prediction (Methods section ‘Prediction validation’), producing a set of confusion matrices for varying subgroups of the validation set (Fig. S2, A to E).

Since the ABNN was trained to functionally reconstruct human physiology and human pathogenesis (Methods section ‘Model training’), its disease-agnostic performance was first assessed for the full validation set (157 predictions; Data S1). The ABNN achieved 90.1% prospective prediction sensitivity (99% CI 80.0%, 96.9%; P < 0.001; Fig. 3B) and 0.86 F1 score (99% CI 0.79, 0.92; P < 0.0001; Fig. 3C; Table 1). It also achieved 82.0% prospective prediction precision (99% CI 70.8%, 90.7%; P < 0.0001; Fig. 3, D and E), a 10.4-fold improvement over the realistic baseline precision of 7.9%, and a 5.4-fold improvement over the conservative baseline precision of 15.1% (Fig. 3D; Methods section ‘Baseline performance’). Additionally, it achieved 82.8% accuracy (99% CI 74.2%, 89.8%; P < 0.0001; Fig. 3C; Table 1) and 72.7% specificity (99% CI 57.5%, 85.3%; P < 0.0001; Fig. 3B; Table 1). These data clearly demonstrate that the ABNN has extracted large-scale, high-quality knowledge, enabling excellent disease-agnostic performance in the real-world settings of PROTOCOLS validation test.

Further analysis of subgroups was conducted for the validation set, with varying degrees of clinical data availability. In theory, the ABNN should not require access to any clinical data prior to making predictions of clinical outcomes for investigational drug candidates (Fig. 2D).

First-in-class clinical trials for novel drug-disease pairs represent a fairly challenging subset of the PROTOCOLS validation test because no prior clinical data are available for these drug-disease pairs across all therapeutic areas. Since PROTOCOLS included 128 first-in-class clinical trials with clinical readouts, the minimum sample and event size requirements for a 99% confidence interval (CI) have been met. The ABNN achieved 92.4% prospective prediction sensitivity (99% CI 80.8%, 99.2%; P < 0.009; Fig. 3, B and F; Table 1), 0.85 F1 score (99% CI 0.76, 0.92; P < 0.0001; Fig. 3C; Table 1) and 78.2% prospective prediction precision (99% CI 64.8%, 88.8%; P < 0.0001; Fig. 3E; Table 1). In addition, it also achieved 82.5% accuracy (99% CI 72.8%, 90.3%; P < 0.0001; Fig. 3C; Table 1) and 71.7% specificity (99% CI 55.6%, 85.0%; P < 0.0001; Fig. 3B; Table 1).

Notably, the ABNN exhibited superior disease-agnostic prospective prediction sensitivity for first-in-class clinical trials, compared with all clinical trials (Δ = 2.3%; 99% CI -0.92%, 5.52%; P < 0.002 for superiority at a 2% margin; Fig. 3F; Methods section ‘Statistical analysis’). The accuracy for first-in-class clinical trials was also non-inferior to its performance for all clinical trials (Δ = 0.3%; 99% CI -0.87%, 1.47%; P < 0.0001 for non-inferiority at a 2% margin; Methods section ‘Statistical analysis’). These results clearly demonstrate that the outstanding performance of the ABNN for PROTOCOLS validation was independent of clinical data availability for arbitrary drug-disease pairs.

Clinical trials in oncology represent a unique challenge for the ABNN, as they conventionally suffer from the lowest clinical success rates. The realistic baseline precision for oncology is 5.3%, in contrast to the highest precision of 23.9% for hematology(*13*) (Methods section ‘Baseline performance’). Consequentially, the importance of preferred positive instances is even higher in oncology than in other therapeutic areas, to the reasonable extent that negative instances are no longer interesting. As such, a clinically meaningful assessment of ABNN performance in oncology should primarily focus on sensitivity, precision, F1 score, and accuracy.

In addition, clinical trials in oncology often include complex trial designs involving active controls and combination therapies as a standard design for pivotal clinical trials. Human cancer patients are further stratified by certain biomarkers (*24*), stages of cancer, and prior treatment history, all of which add to the complexity of scientific knowledge required to make accurate predictions of clinical drug efficacy and safety.

The PROTOCOLS validation set included 48 clinical trials in oncology with clinical readouts, satisfying the minimum sample and event size requirements for a 90% CI have been met (see Methods section ‘Statistical analysis’). The ABNN achieved 88.9% prospective prediction sensitivity (90% CI 75.7%, 94.3%; P < 0.015; Fig. 3B; Table 1) and 0.85 F1 score (90% CI 0.77, 0.90; P < 0.0004; Fig. 3C; Table 1). Remarkably, the ABNN achieved 82.1% prospective prediction precision (90% CI 68.9%, 89.3%; P < 0.003; Fig. 3, B and G; Table 1), a 15.5-fold improvement over the realistic baseline precision of 5.3%, and a 7.6-fold improvement over the conservative baseline precision of 10.8% for oncology clinical trials (Methods section ‘Baseline performance’). It also achieved 77.1% prospective prediction accuracy (90% CI 65.1%, 84.9%; P < 0.0004; Fig. 3C; Table 1). These data clearly demonstrate the ABNN has extracted sufficiently complex and intricately nuanced knowledge of the human body and cancer etiology to accurately predict the clinical success of cancer drugs in biomarker-differentiated heterogeneous patient populations.

COVID-19 represents the ultimate test of existing knowledge quality for the ABNN because no knowledge of COVID-19 can be extracted directly, due to a lack of prior data shortly after the outbreak. As such, the entire disease model for COVID-19 can only be constructed by generalizing other knowledge to COVID-19 clinical symptoms. In addition, no clinical data were available worldwide when the preprint of the ABNN COVID-19 disease model was published on March 20, 2020 (*25*).

As there are 49 infectious disease clinical trials in the validation set, the minimum sample size and event size requirements for a 90% CI have been met (see Methods section ‘Statistical analysis’). In total, 94% (46/49) are pivotal COVID-19 clinical trials (Data S1, T4). The ABNN achieved 88.9% prospective prediction sensitivity (90% CI 68.3%, 95.4%; P < 0.05; Fig. 3G; Table 1) and 0.78 F1 score (90% CI 0.65, 0.86; P < 0.0012; Fig. 3C; Table 1).

The ABNN also achieved 69.6% prospective prediction precision (90% CI 51.8%, 81.6%; P < 0.0035; Fig. 3G; Table 1) for infectious diseases in the validation test. This represents a 5.3-fold improvement over the realistic baseline precision of 13.2% and a 3.1-fold improvement over the conservative baseline precision of 22.8% for infectious disease clinical trials (Methods section ‘Baseline performance’). Additionally, it also achieved 81.6% accuracy (90% CI 70.1%, 88.4%; P < 0.002; Fig. 3C; Table 1). These data clearly indicate the quality of extracted knowledge in the ABNN has reached a critical mass, enabling *de novo* generation of comprehensive knowledge for a new disease like COVID-19 while maintaining the same quality as that of the new knowledge extracted from prior biological data.

The ABNN achieved 92.4% ± 7.8% prospective prediction sensitivity (recall), 0.91 ± 0.10 F1 score (Fig. 3C; Table S1), and 90.0% ± 12.0% prospective prediction precision (Fig. 3, B and E; Table S1), across all other therapeutic areas in the PROTOCOLS validation set (60 predictions in total; Data S1, T7a to T7d). Although the minimum requirements for a 90% CI were not met for these data (Methods section ‘Statistical analysis’), the results demonstrate that ABNN performance is stably high and consistent across therapeutic areas.

Alzheimer’s disease (AD) was also included in the PROTOCOLS validation test and served to further demonstrate the robustness of extracted knowledge in the ABNN. AD represents an extreme case because the historical clinical success rate of disease-modifying AD drugs is nearly zero (*26*). Sensitivity, or even a non-zero true positive, thus becomes the most appropriate performance metric for AD because both false positive and false negative instances are equally uninteresting in real-world settings. As such, AD represents a significant challenge for the ABNN because of a consensus that it is overly complex, the quantity of existing data is too low, and the quality of existing data is poor (*26*).

Despite these limitations, the ABNN achieved 100% sensitivity and 100% precision with non-zero true positives and no false positives (Data S1, T6) for three prospective predictions of AD pivotal clinical trials during PROTOCOLS validation. The first true positive was Masitinib, a first-in-class c-Kit inhibitor that met the efficacy primary endpoint in a pivotal phase 3 trial (NCT01872598). These results demonstrate that the ABNN has extracted large-scale, high-quality knowledge that warrants a thorough understanding of AD etiology from existing data previously believed to be insufficient.

Aging is an evolutionarily conserved effect across all mammalian species (*27*) and is clinically recognized as the primary risk factor for numerous diseases across multiple therapeutic areas (*28*). The quality of extracted aging-specific knowledge in the ABNN was evaluated by identifying all age-related pivotal clinical trials in the PROTOCOLS validation test (Methods section ‘Validation data collection’) and corresponding ABNN performance (Fig. 4, A and B).

**Fig. 4.**
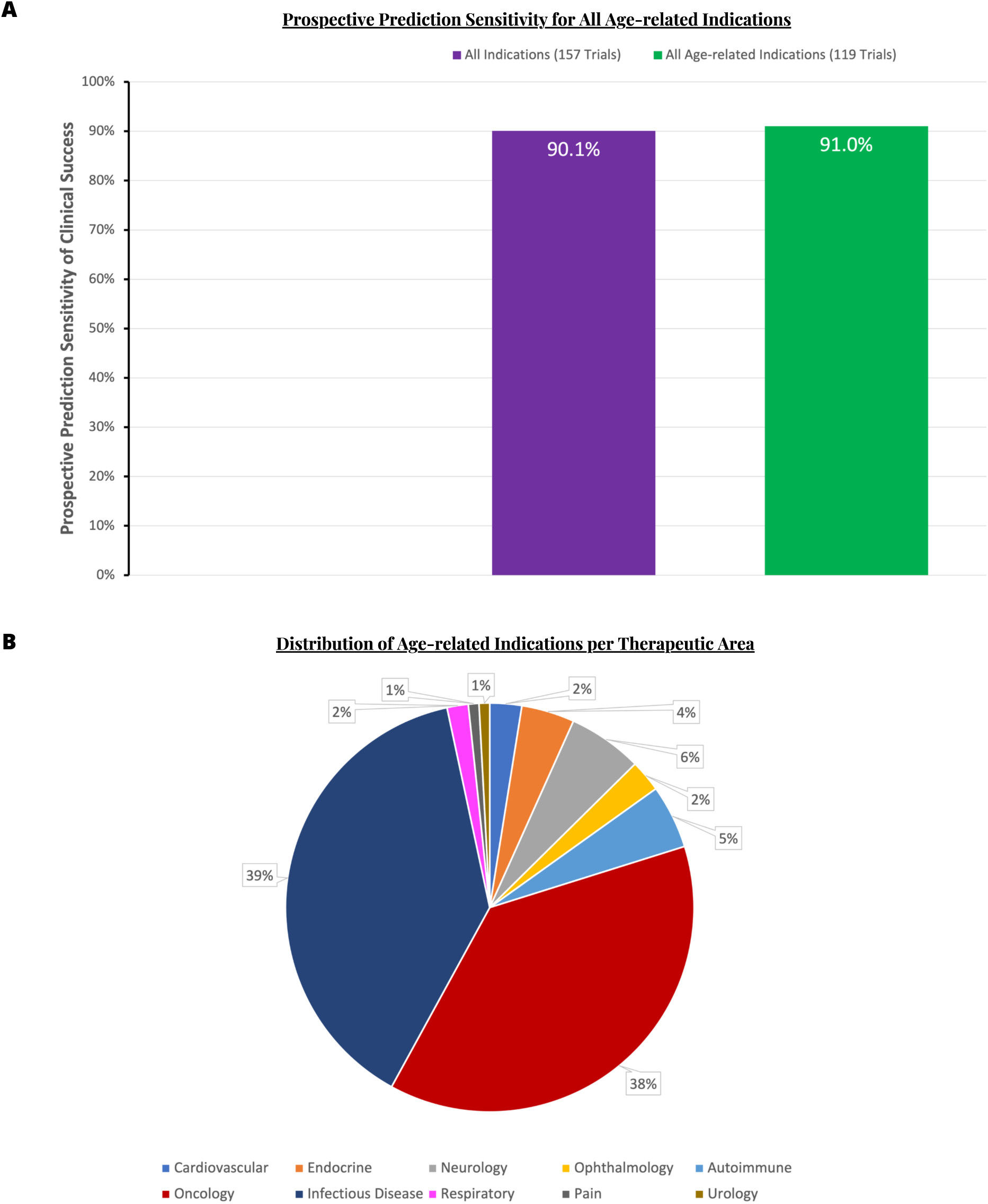
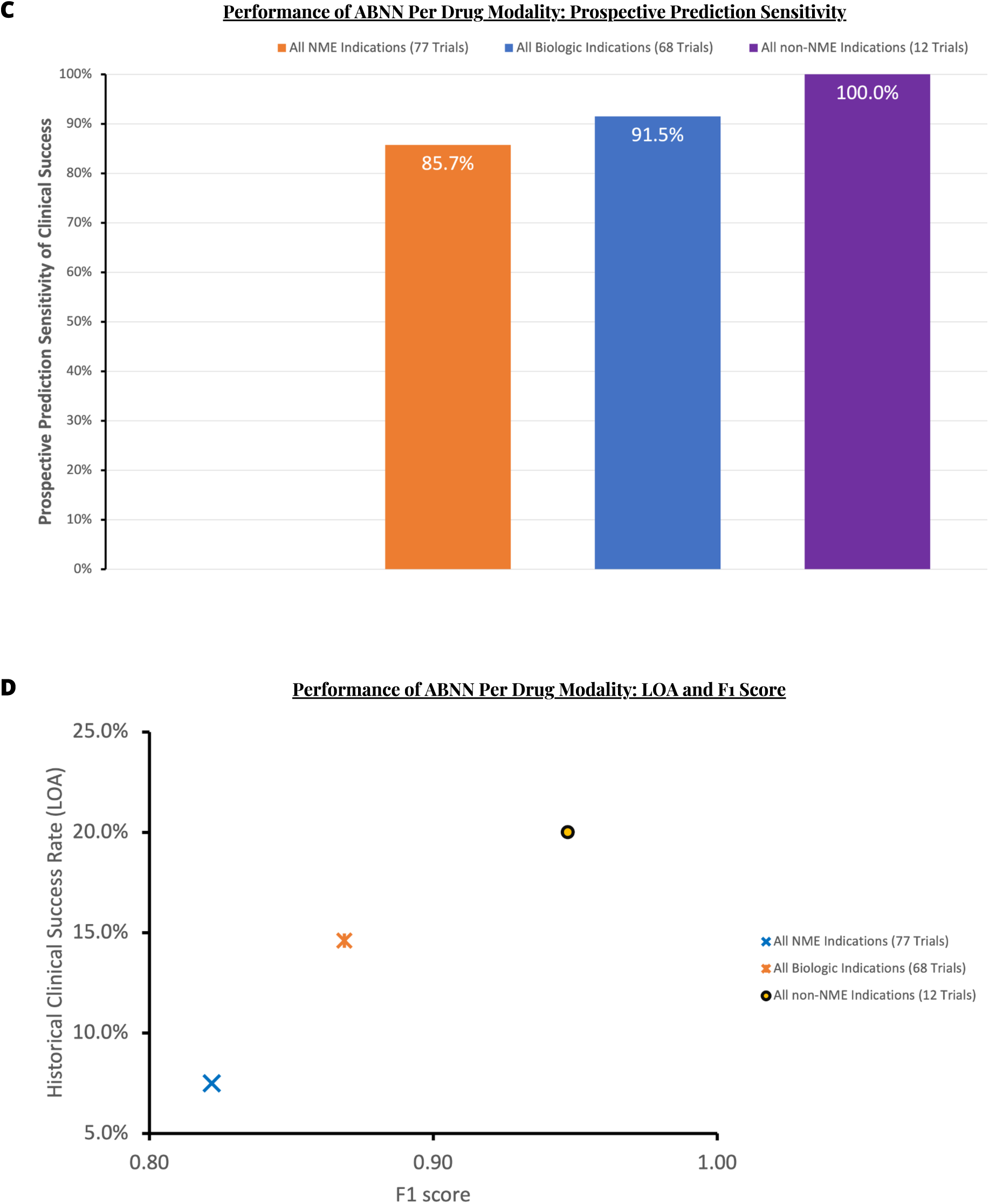
Prospective prediction performance metrics across age-related diseases and drug modality. (**A**) Prospective prediction sensitivity of all age-related pivotal clinical trials was 91.0% (99% CI 79.1%, 98.4%; P < 0.005) in the PROTOCOLS validation test (Methods section ’Statistical analysis’). (**B**) The distribution of all age-related pivotal clinical trials per therapeutic area in the PROTOCOLS validation set (Methods section ’Validation data collection’). (**C**) ABNN performance as measured by prospective prediction sensitivity across drug modalities (see Data Table 1 for more details). ABNN achieved superior performance for biologics than for small molecules (Δ = 5.8%; 90% CI 0.78%, 10.82%; P < 0.0001 for superiority at a 2% margin; Methods section ‘Statistical analysis’). (**D**) ABNN performance as measured by historical clinical success rates for investigation new drugs (LOA) and F1 scores (see Table 1 for details). LOA is identical to the realistic baseline accuracy (see Methods section ’Baseline performance’). F1 scores and LOAs across overall validation subgroups are strongly positively correlated (exact Pearson coefficient = 0.975).

As there are 119 age-related clinical trials of age-related diseases in the validation test, the minimum sample and event size requirements for a 99% CI have been met. The ABNN achieved 91.0% prospective prediction sensitivity (99% CI 79.1%, 98.4%; P < 0.005; Fig. 4A; Table 1) and 0.85 F1 score (99% CI 0.76, 0.92; P < 0.0001; Fig. 3C; Table 1). It also achieved 79.2% prospective prediction precision (99% CI 65.9%, 89.7%; P < 0.0001; Fig. 3E; Table 1), 81.5% accuracy (99% CI 71.3%, 89.7%; P < 0.0001; Fig. 3C; Table 1), and 69.2% specificity (99% CI 51.8%, 83.9%; P < 0.0001; Fig. 3B; Table 1).

Remarkably, ABNN prospective prediction sensitivity for all age-related clinical trials was non-inferior to its performance for all first-in-class clinical trials (Δ = 1.4%; 99% CI -1.12%, 3.92%; P < 0.003 for superiority at a 2% margin; Methods section ‘Statistical analysis’). ABNN prospective predictive accuracy for age-related clinical trials was also shown to be non-inferior to its performance for first-in-class clinical trials (Δ = 1.0%; 99% CI -1.14%, 3.14%; P < 0.0001 for non-inferiority at a 2% margin; Methods section ‘Statistical analysis’).

ABNN prospective predictive sensitivity for all age-related clinical trials was statistically significantly higher than its performance for all clinical trials in the validation set (Δ = 0.9%; 99% CI -1.13%, 2.93%; P < 0.0001 for superiority at a 2% margin; Fig. 4A; Methods section ‘Statistical analysis’). These results clearly demonstrate the ABNN has extracted sufficient knowledge of aging, which contributed significantly to its consistent performance across all the age-related indications in the PROTOCOLS validation set (Fig. 4B).

We also investigated the relationship between the quality of extracted knowledge in the ABNN and drug modality (Fig. 4C). The ABNN achieved 91.5% prospective prediction sensitivity (90% CI 80.8%, 95.7%; P < 0.015; Fig. 4C; Table 1) for biologics (68 biological trials; Methods section ‘Drug classification’) and 85.7% prospective prediction sensitivity (90% CI 71.9%, 92.2%; P < 0.009; Fig. 4C; Table 1) for small molecule drugs (77 NME trials; Methods section ‘Drug classification’). This performance was consistently high across drug modalities and even higher for biologics than for small molecules (Δ = 5.8%; 90% CI 0.78%, 10.82%; P < 0.0001 for superiority at a 2% margin; Fig. 4C; Methods section ‘Statistical analysis’). We further calculated correlations between F1 scores and realistic baseline precisions (or LOA: historical clinical success rates for investigational new drugs since phase 1. See Methods section ‘baseline performance’) across drug modalities. F1 scores and LOA were positively and strongly correlated (Exact Pearson r = 0.975; Fig. 4D), indicating that the quality of extracted knowledge in the ABNN may be a natural fit for drug modalities with a higher target specificity.

Finally, we investigated the relationship between the quality of extracted knowledge in the ABNN and the difficulty of drug discovery per therapeutic area, by calculating correlations between F1 scores and realistic baseline precisions (or LOA: historical clinical success rates for investigational new drugs since phase 1; see Methods section ‘baseline performance’) for all validation subgroups (Fig. 5A). F1 scores and LOA were negatively and weakly correlated (Exact Pearson r = -0.260), indicating that the quality of ABNN knowledge concerning human pathogenesis is orthogonal to the quality of collective knowledge for corresponding therapeutic areas. Surprisingly, ABNN performance was higher for increasingly difficult therapeutic areas (Fig. 5A; Table 1). This observation suggests that the ABNN is optimized for modelling defining characteristics of systematic diseases that are less amenable to the divide-and-conquer approaches that work well for local diseases. This is consistent with another surprising finding that ABNN prospective prediction sensitivity (recall) and ABNN prospective prediction precision are strongly and positively correlated across validation subgroups (Exact Pearson r = 0.808; Fig. 3E), in stark contrast to the typical inverse relationship between precision and recall for binary classifications (*20*). This finding reveals that ABNN performance is primarily driven by improved disease understanding that unbiasedly reduces both false positives and false negatives, rather than by biased cutoff tuning that trades off false positives against false negatives.

**Fig. 5.**
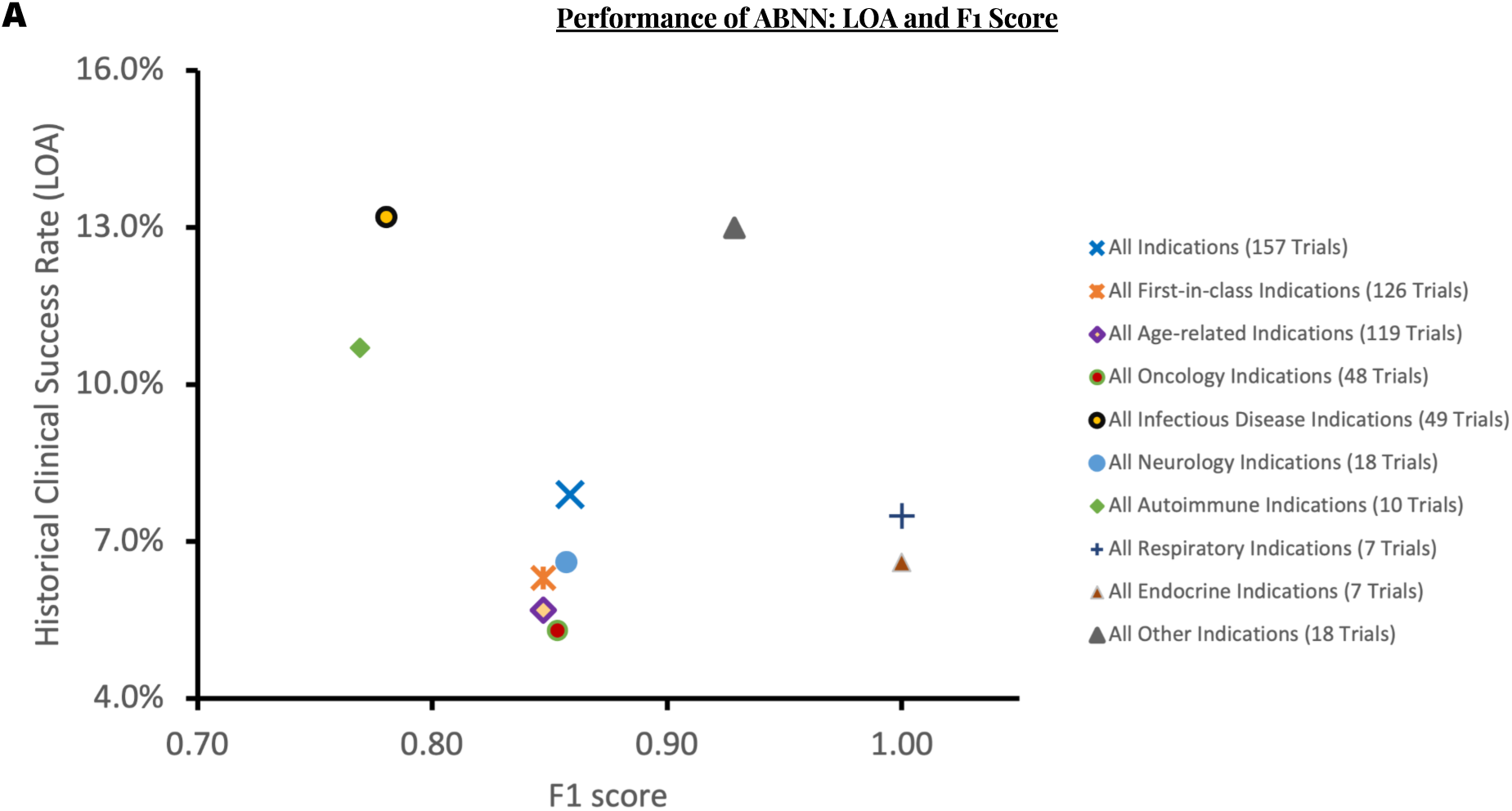
Better performance for harder diseases. (**A**) ABNN performance as measured by historical clinical success rates for investigation new drugs (LOA) and F1 scores (see Table 1 for details). LOA is identical to the realistic baseline precision (see Methods section ’Baseline performance’). F1 scores and LOAs across overall validation subgroups were negatively and weakly correlated (exact Pearson coefficient = -0.260).

## DISCUSSION

In this study, the possibility of a deep-learning-augmented human intelligence has been presented for constructing a foundation model of human physiology and human pathogenesis, potentially enabling highly accurate data-free predictions of clinical success for investigational new drugs applied to almost any disease. This achievement is intractable for clinical-data-driven machine intelligence, on which the demands for clinical data are unrealistically high and the efficiency of knowledge extraction is sufficiently low that gaps between promise and proof have become nearly unbridgeable (Fig. 1).

Humans have designed machine learning methods to train deep neural networks that have successfully learned black-box models, outperforming human experts in playing games (*1*) and predicting protein structures (*2*). Our work shows that a similar approach could be adapted to training augmented biological neural networks (ABNNs) in learning quasi-white-box models (*25*) for predicting the clinical success of investigational new drugs (Fig. 1B).

The resulting ABNN could safely deliver AI-surpassing clinical utility while averting the privacy and trustworthiness challenges faced by medical AI systems (*16*). The ABNN could also improve the design of labels for training better-performing DNNs with fewer data (*29*).

Furthermore, in contrast to domain experts who have privileged access to a large body of private preclinical data, the presented ABNN was trained solely using public preclinical data yet extracted more knowledge than previous attempts by experts (Fig. 2D).

PROTOCOLS represents the most rigorous validation test to date for evaluating the real-world utility of extracted knowledge from large-scale, low-quality data spaces (Fig. 2, and Fig. 3A). With a 99% CI, the trained ABNN achieved a prospective prediction sensitivity of more than 90%, an F1 score of more than 0.85, a prospective prediction precision of more than 80%, and a prospective prediction accuracy of more than 82% across all therapeutic areas, independent of the availability of prior clinical data (Table 1).

In addition, the drug mechanisms of action and clinical trial design protocols were the only inputs used by the ABNN to evaluate the clinical success of novel drugs (Fig. 2D). ABNN performance in PROTOCOLS confirmed that large-scale, high-quality scientific knowledge alone may be sufficient to accurately predict drug clinical success.

Given that both ABNN inputs are available prior to first-in-human studies, the ABNN could be integrated into standard workflows of clinical development as an add-on screening of shortlisted drug candidates, to maximize clinical benefit and clinical success for investigational new drugs. The ABNN could not only discover 90.1% of all possible drug-indication pairs that could bring clinical benefits to patients, but also increase tenfold the baseline clinical success rate from 7.9% to 82.0%, ushering a permanent reversal of Eroom’s law (Fig. 2D) (*4*).

The validated ABNN is also the world’s first clinically validated model of human physiological aging and age-related diseases (Fig. 4, A and B). In parallel with improving the productivity of developing disease-specific therapeutics, it provides a feasible path for developing disease-agnostic medicines that safely and precisely target underlying mechanisms of aging to potentially delay or prevent the onset of age-related diseases (*30*). This could significantly ease the escalating burden of aging populations on global healthcare systems.

The ABNN also leaves room for improvement as 17.2% (27/157) of predictions were either false positives (18/157) or false negatives (9/157) in the PROTOCOLS validation. The ABNN is also limited to model a small fraction of rare disorders and hematological disorders (Methods section ‘Validation data collection’). As such, further work is planned to investigate why the ABNN performs better for more difficult diseases (Fig. 5A). The exponential efficiency of knowledge extraction enabled the ABNN to translate each false prediction into substantial improvements in the corresponding disease model.

Another potential limitation of our work is that the ABNN was validated in PROTOCOLS test using ongoing pivotal clinical trials whose drug candidates had already been pre-selected by experts to enter clinical development. ABNN performance in development candidate selection may thus be conditioned upon prior expert contributions, the extent of which could be determined in future rounds of PROTOCOLS validation. This could be enabled by an expanded access to private preclinical data, allowing for the ABNN to predict the clinical success of drug candidates ruled out by experts. ABNN predictions could thus be used to initiate clinical-stage development for drugs with high clinical success potential that would have otherwise been dismissed.

Regardless, the possible conditioning of ABNN performance on prior expert performance in development candidate selection should be seen as a real-world advantage in application. The ABNN enables a seamless integration with any standard workflow in clinical development as an add-on screening of expert-shortlisted development candidates, without making significant modifications to standard procedures. As such, the model serves as an augmented intelligence in collaboration with experts, in contrast to other artificial intelligence systems that are often positioned in competition with or even in substitution for experts (*16*).

While this work could significantly reshape medicine, its broader impact lies in the presented training methodology, which provides a new path for drastically increasing the efficiency of knowledge extraction in every discipline that is characterized by large-scale, low-quality data spaces (e.g., synthetic biology, material science, and climate science). These results also provide the first prospective proof that the smallest research team is necessary for delivering the most disruptive breakthroughs by enabling the largest ABNN optimized for discovering novel knowledge for complex challenges (*31*).

The advent of the ABNN represents a new era of human-centric augmented intelligence. The global adoption of ABNN training could shift research culture and organizational structure towards a potential mass production of scientific and technological breakthroughs. ABNNs could give rise to general purpose technologies that may potentially overcome the productivity J-curve and paradoxes recurrently observed in the past (*32*).

## Data Availability

All data produced in the present study are available upon reasonable request to the authors.

## ACKNOWLEDGMENTS

To be added.

## AUTHOR CONTRIBUTIONS

Conceptualization: BL, IL

Methodology: BL, IL

Investigation: BL, IL

Visualization: BL, IL

Validation: BL, IL

Formal analysis: BL

Resources: IL

Funding acquisition: IL

Project administration: IL, BL

Supervision: BL, IL

Writing – original draft: BL

Writing – review & editing: BL, IL

## DATA AVAILABILITY

The PROTOCOLS validation raw data are publicly available at twitter.com/demiurgetech.

All data are available in the main text or the supplementary materials.

## Supplementary Materials

### Materials and Methods

#### TRAINING DATA COLLECTION

Two types of training data were collected for different stages of ABNN training.

In stage 1, the ABNN was trained to learn a foundation model of the normal states of the human body to capture the latent complexity of human physiology. The stage 1 training data covered all publicly accessible sources of biological data that could inform normal physiological processes in the human body, including cross-species data about evolutionarily conserved biological building blocks (Methods section ‘Model training’).

In stage 2, the ABNN was trained to learn disease-specific models of the abnormal states of the human body, which should match the clinical scope of human pathogenesis. The stage 2 training data covered all publicly accessible sources of biological data that investigated disease etiology in cell cultures and animal models, as well as medical data reporting disease-specific co-morbidities and symptoms in human patients. However, no clinical trial data were collected for ABNN training because they were designed for drawing causal inferences about drug-body interactions under an assumption that the human body is a black-box system, thus uninformative for modelling the underlying mechanisms of human pathogenesis (Methods section ‘Model training’).

#### MODEL TRAINING

Inspired by the multi-faceted functional correspondence between deep neural networks (DNNs) and biological neural networks (BNNs) (*7*), we adapt the best practice of training DNNs into a standard protocol of training BNNs to learn representations from large-scale data spaces like DNNs, resulting in augmented biological neural networks (ABNNs) as a form of replicable augmented intelligence. We use our validated ABNN in life science as an example for illustrating each step in the training protocol:

##### Training Data

DNNs learn better representations of data with more levels of abstraction than with fewer levels, and BNNs share a hierarchical architecture with DNNs (*7*). Accordingly, ABNNs must be exposed to the full breadth and depth of multi-omics data (e.g., genome, proteome, transcriptome, epigenome, metabolome, and microbiome) to capture the latent complexity of human physiology and the clinical scope of human pathogenesis (Fig. S1A). More specifically, the ABNN extracted right knowledge by being exposed to the entire English corpus of non-clinical-trial biomedical publications in journals whose impact factors were stably greater than two over the three most recent consecutive years (including but not limited to: Science, Nature, Cell, Nature Neuroscience, Nature Metabolism, Cell Stem Cell, Neuron, PNAS, eLife, Nature Communication, Scientific Reports, etc.), as well as public repositories of curated databases (including but not limited to: Human Protein Atlas, Allen Human Brain Atlas, The Cancer Genome Atlas, etc.).

##### Learning Rules

DNNs use backpropagation to improve learned representations of data by sequentially updating its internal parameters from the highest to the lowest level of abstraction. Ubiquitous feedback connections in BNNs can then be used to implement backpropagation-like learning rules (Fig. S1B). ABNNs extract fast knowledge by iteratively updating internal representations in a strict order from the most macroscopic levels (phenotype) through intermediate levels (endophenotype) to the most microscopic levels (genotype). The ABNN was trained to focus on a single area of interest across all levels of abstraction until backpropagation-like learning was completed for each area before moving on to the next. For example, when cortical columns used for motor control were the training area of interest, the ABNN translated every piece of training data into a sequential update to internal representations in the ascending order of granularity from the phenotype level (e.g., muscle contractions), to the endophenotype level (neuronal circuits responsible for muscle contractions), to the cell level (e.g., Layer 5 pyramidal neurons (L5PT) and Layer 2/3 inhibitory interneurons (L2/3IN) responsible for muscle contractions), to the protein level (e.g., NMDA receptor subtypes and GABA receptor subtypes specifically expressed in L5PT and L2/3IN for muscle contractions), to the epigenotype level (e.g., DNA methylation profiles in connection with subtype-specific NMDAR and GABAR activities in L5PT and L2/3IN for muscle contractions), and finally to the genotype level (e.g., the potential role of the CLOCK gene expression on subtype-specific NMDAR and GABAR activities in L5PT and L2/3IN for muscle contractions).

##### Learning Rate and Loss

DNNs learn general-purpose representations of all-encompassing data to deliver optimal performance in a wide range of special-purpose tasks (*11*). Both BNNs and DNNs can learn to command general linguistic capabilities for numerous tasks. ABNNs extract full-scope knowledge by first learning a general-purpose model of human physiology from the all-encompassing non-disease biological data (Stage 1; Methods section ‘Training data collection’), prior to learning human disease models from disease-specific data (Stage 2; Methods section ‘Training data collection’) (Fig. S1C). In Stage 1, small learning rates and losses were used so the ABNN was incentivized to conduct multiple rounds of learning without being sensitive to the learning performance of general human physiology in each round. In Stage 2, the learning rate and loss were set to higher values so that the ABNN would be incentivized to learn a complete model of a specific human disease in a single round. The ABNN was thus incentivized to re-run Stage 1whenever performance was suboptimal in Stage 2. As a result, the ABNN optimized the global extrema of the disease-agnostic physiology model before optimizing the local extrema of any disease-specific pathogenesis model. This two-stage learning has been crucial for enabling the de novo model generation for new diseases like COVID-19 (*33*).

##### Network Size

DNNs maximize the robustness of learned representations of data by increasing the sheer size of network parameters (*10*). ABNNs extract novel knowledge if a single ABNN of the largest size learns from the entirety of the all-encompassing data, as opposed to multiple smaller ABNNs learning from siloed sub-datasets (Fig. S1D). In practice, this means that we should minimize the number of ABNNs in a research team to one, to maximize the number of ABNN neuronal nodes interconnected by always-on high-bandwidth biological synapses in a single brain, rather than by the intermittently-on low-bandwidth human languages between several brains.

##### Training Environment

DNNs learn the best representations of data when sufficient time and ample resources allow for an uninterrupted pass of the full training data, prior to which no meaningful performance milestones should be defined (*9*). As such, the ABNN was not evaluated before the completion of learning from both human physiology and human diseases (Fig. S1E), no matter how long it would take the ABNN to complete learning. Securing an optimal environment was the most challenging aspect of ABNN training, as it would be extremely difficult to establish a research culture and implement terms and protocols for uninterrupted training that may last for a decade without intermediate milestones.

##### Overfitting Prevention

The ABNN may be immune to overfitting because it is sufficiently generalizable for learning from data that are categorically heterogeneous for the intended application. In contrast, overfitting is somewhat inevitable with a DNN because it lacks this generalizability and cannot learn from data that are not perfectly homogeneous for an intended application (Fig. 1B). In the context of life science and drug discovery, the ABNN was first trained using biological data reflecting normal physiological states, and then tested solely using clinical data representing abnormal pathological states. The ABNN refrained from accessing disease-specific clinical data until and unless the etiology of a specific disease was entirely generated de novo from the foundation model of human physiology in the first place. The resulting ABNN should be able to directly extrapolate from the normal states of human physiology to the abnormal states of human diseases.

#### VALIDATION DESIGN AND METHODOLOGY

Validation is critical for an objective assessment of ABNN capabilities in exponentially extracting high-quality knowledge from life science data (Fig. 2, A and B). The default approach is to determine the extent to which the ABNN could reverse Eroom’s law. However, it would require a multi-billion-dollar budget and a multi-decade timeline to have at least 118 new clinical-stage drug candidates from scratch to assess ABNN’s capabilities of disease-agnostic high-fidelity body-drug interactions with a statistical significance of 99%. The default validation approach is thus unrealistically time-consuming and non-scalable. As such, running virtual clinical trials is an effective alternative. Drug mechanisms of action and clinical trial design protocols would serve as the only inputs and the ABNN could be used to predict the clinical efficacy and safety of new drugs solely using extracted knowledge, without access to patient data or confidential third-party information. Highly accurate virtual clinical trials could be run before first-in-human studies to prevent drug developers from initiating pivotal real-world trials that are doomed to fail (*34*).

A rigorous validation of the ABNN, including the stringent criteria from the PROTOCOLS test (Fig. 2B), would involve publishing prospective predictions for pivotal clinical trial outcomes across all major human diseases. Tamper-proof timestamps would be included and prediction precision could be determined with a 99% CI, based on a ground truth determined by actual clinical readouts. No selection of lead over non-lead indications would be included, although non-lead indications have a far lower success rate (*14*).

There are a few hundred new pivotal clinical trials each year whose protocols are publicly accessible in a centralized database (as required by the FDA (*35*)) and whose clinical readouts are typically published by sponsors in a standardized format. Public, prospective predictions of pivotal ongoing clinical trial outcomes could serve as a benchmark validation for the ABNN, similar to the way in which ImageNet provided a benchmark validation for visual object recognition using DNNs (*36*).

#### METRICS DEFINITION

Confusion matrices were produced to assess ABNN performance on a disease-agnostic basis and are shown per therapeutic-area basis (Fig. S2, A to E). The following metrics are defined and applied to true positives (TP), true negatives (TN), false positives (FP), and false negatives (FN) in those confusion matrices:

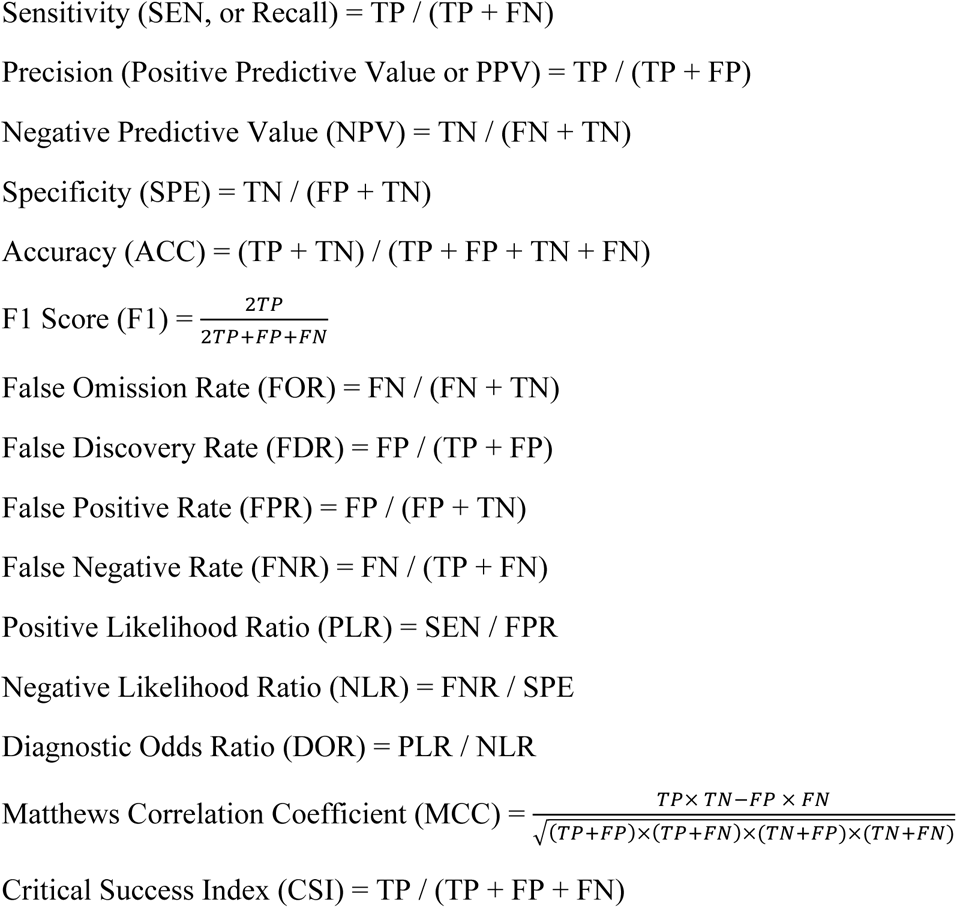

#### BASELINE PERFORMANCE

Prospective prediction precision (PPP) is defined as the percentage of clinical programs that have achieved actual clinical success by demonstrating sufficient clinical efficacy and safety in pivotal clinical trials that warrant an FDA approval, given that clinical programs are initiated only for those drug candidates that are prospectively predicted to be clinically successful by experts.

Realistic baseline precision (RBP), as a realistic estimate of PPP, is defined as the probability of FDA approval for drugs in phase 1 development (Phase1 likelihood of approval, or P1LOA). It was calculated using corresponding P1LOAs in a 2011-2020 survey of historical clinical trials (*13*). This baseline precision is realistic as it acknowledges that the ABNN would make identical predictions at earlier stages of development with drug mechanisms of action and clinical trial design protocols serving as the only inputs, both of which are available as early as the preclinical stages.

Conservative baseline precision (CBP), as a conservative estimate of PPP, is defined as the probability of FDA approval for drugs in phase 2 development (Phase2 likelihood of approval, or P2LOA). It was calculated as the corresponding P2LOA in a 2011-2020 survey of historical clinical trials (*13*). This baseline precision is conservative because it does not consider that the ABNN only requires two inputs to generate predictions (drug mechanisms of action and clinical trial design protocols, but not phase 1 clinical data), both of which are readily available in the real world even before first-in-human studies (phase 1). Regardless, this baseline precision is more balanced in consideration of both the phase distribution for clinical trials in the validation set (157 clinical readouts (5/157 = 3.18% phase 1 trials, 69/157 = 43.9% phase 2 trials, and 85/157 = 54.1% phase 3 trials, and the irrelevance of phase transitions in making predictions.

RBP was not directly available from previous survey studies (*13, 14*) for two subgroups of the PROTOCOLS validation set: all first-in-class indications (128 trials) and all age-related indications (119 trials). As diseases in these two subgroups originate from therapeutic areas featuring lower-than-average historical clinical success rates (oncology, neurology, cardiovascular, etc.), the real RBPs for both subgroups should be lower than those of all indications (7.9%) (*13*). We therefore assumed that a reasonable estimate of RBP for the subgroup containing all first-in-class indications (128 trials) would be 6.32%, equal to 80% of the RBP for all indications (7.9%) (*13*). Since no anti-aging therapeutics have ever been clinically approved, we assumed that a reasonable estimate of RBP for the subgroup of all age-related indications (119 trials) would be 5.69%, equal to 72% of the RBP for all indications (7.9%) (*12*).

#### VALIDATION DATA COLLECTION

After completing the two-stage training, the ABNN functions as a ‘virtual patient’ that has functionally reconstructed both human physiology and human pathogenesis with sufficient fidelity. The trained ABNN is thus clinically equivalent to a real human body such that ‘virtual clinical trials’ could be run on the ABNN to evaluate realistic drug-body interactions and predict clinical success in human patients. Clinical trial data were thus all reserved for testing the fully trained ABNN in the real-world PROTOCOLS validation test.

The validation test comprises the registration information for human clinical trials on www.clinicaltrials.gov, with the screening criteria as shown in Fig. 2C (phase 1/2 is categorized as phase 1, and phase 2/3 is categorized as phase 2). We estimated that there are approximately 200-300 clinical trials each year that would meet these screening criteria, which is consistent with the 424 annual clinical trials exhibiting a much looser set of screening criteria (no limit on primary completion date, no preference of first-in-class trials, etc.) in a comprehensive survey study (*14*). See Data S1 for the full raw data of PROTOCOLS test .

A clinical trial in the validation set was designated as “first-in-class” if the drug-indication pair had not been approved for marketing worldwide.

A clinical trial in the validation set was designated as “age-related” if age was a clinically identified risk factor for the indication thereof.

The ABNN is principally disease-agnostic yet leaves room for improvements in identifying hematological disorders, rare neurodevelopmental disorders, and rare musculoskeletal disorders. These unfinished disorders were excluded from the performance validation step yet were included in validation data collection because prospective predictions can provide additional insights to accelerate model building for such disorders.

#### PREDICTION GENERATION

Per industry-wise best practices (*37*), the ABNN predicted SUCCESS for a pivotal clinical trial if at least one of its pre-specified efficacy primary endpoint(s) was believed to be met with predefined statistical significance. In contrast, the ABNN predicted FAILURE for a pivotal clinical trial if none of its pre-specified efficacy primary endpoint(s) were believed to be met with predefined statistical significance (Fig. 2D).

If a pivotal clinical trial had multiple co-primary endpoints, PARTIAL SUCCESS or PARTIAL FAILURE was predicted if at least one of its pre-specified efficacy primary endpoint(s) was believed to be met with predefined statistical significance and at least one of its prespecified efficacy primary endpoint(s) was believed to not be met with predefined statistical significance.

#### PREDICTION PUBLICATION

A total of 265 predictions were published as timestamped tweets @DemiurgeTech to establish a publicly accessible track record for prospective predictions (Fig. S3). The immutability of tweets ensures that each published prediction could not be post hoc modified. We also indexed each tweet to prevent any intentional deletions of false predictions that go unnoticed, to ensure a reliable performance evaluation. However, index gaps do exist for several legitimate cases, which does not compromise the rigor of validation for the following reasons. 1) An index was unintentionally skipped and remained unused from that point forward. 2) An indexed prediction was made but later became disqualified as a prospective prediction, because the corresponding result had been announced after the indexed prediction was made but before the indexed prediction was published. We used DocuSign to distinguish these two cases of index gaps by electronically signing every indexed prediction before publishing it as a tweet. As such, no DocuSign certificates are available for unused indices. The indices 001 and 286 denote the indices of the first and the last published prospective predictions, respectively. A total of 21 index gaps were detected, 5 of which were unused and 5 of which were for disqualified predictions lacking clinical readouts as of the cutoff date (March 1, 2022). Nine predictions were made for disqualified predictions with clinical readouts as of the cutoff date (3 false predictions and 6 true predictions). Two predictions were for qualified unpublished predictions whose results are still not available. DocuSign certificates for disqualified and qualified unpublished predictions are available upon written request (Fig. 2D).

#### CLINICAL READOUT

It is customary for private companies and mandatory for public companies in the biopharmaceutical industry to forecast the estimated dates of clinical readouts and to publish the actual results from human clinical trials. We tracked the availability of clinical readouts for every published prospective prediction from multiple online sources, including https://www.globenewswire.com/ for press releases, and https://www.biospace.com/ for curated news, and financial reports from public companies. We published links to available clinical readouts as timestamped tweets by replying to the original timestamped tweet for the corresponding prediction, thus establishing a clear timeline of predictions followed by readouts (Fig. 2D).

#### GROUND-TRUTH DETERMINATION

Under the FDA guidelines (*38*), a clinical trial with available readouts is assigned the ground-truth label *success* if and only if at least one efficacy primary endpoint is met according to interim/topline analyses or final data published by the trial sponsors (Fig. 2D).

Similarly, a clinical trial with available readouts is assigned the ground-truth label *failure* if and only if (a) none of the efficacy primary endpoint(s) are met according to topline analyses or final data or none are deemed likely to be met according to interim analyses; (b) the trial is terminated for non-recruitment or non-business reasons; (c) the trial is terminated after the failure of other clinical trials for identical drug-disease pairs; (d) the post-trial development is discontinued for non-business or unspecified reasons; (e) the trial is dropped out from the pipeline without specific reasons. All of these criteria were determined using official websites or announcements by the trial sponsors.

Given that the reliability of ground-truth determination is capped by an 85.1% theoretical maximum prediction accuracy for drug clinical success in human clinical trials (*39*), we refrained from directly labeling missed predictions as false without further consideration of subsequent developments. Specifically, we considered missed predictions of clinical failure for phase 2b trials that were less statistically powered than phase 3 trials. Missed predictions of clinical failure should thus be more likely to be true rather than false if the trial sponsor decided not to conduct a phase 3 trial for non-business or unspecified reasons.

#### PREDICTION VALIDATION

A prospective prediction of *success* was validated as TRUE if its corresponding ground-truth label was also *success* or invalidated as FALSE if its corresponding ground-truth label was *failure*. Similarly, a prospective prediction of *failure* was validated as TRUE if its corresponding ground-truth label was also *failure* or invalidated as FALSE if its corresponding ground-truth label was *success*. Confusion matrices were produced to assess ABNN validation performance on a disease-agnostic basis and are shown per therapeutic-area basis (Fig. S2, A to E).

#### DRUG CLASSIFICATION

Drugs in the PROTOCOLS validation set were generally categorized using the FDA classification codes for new drug applications: new molecular entity (NME), biologic, and non-NME codes compromising vaccines and other modalities.

NMEs are novel small molecule drugs or novel combinations of multiple old or new small molecule drugs. Biologics comprise a broad range of drug types such as antibodies, peptides, RNAi therapies, cell therapies, gene therapies, and microbiome therapies. Non-NMEs include vaccines, cytokines, enzymes, insulins and other reformulations of approved drugs.

#### STATISTICAL ANALYSIS

ABNN performance in clinical success prediction was evaluated using the public, prospective prediction of pivotal ongoing clinical trial success outcomes at large scale (PROTOCOLS) validation test, a rigorous external validation of a clinical prediction model with a binary outcome (*23*).

Multiple reasonable assumptions were included to calculate minimal sample and event sizes, for the determination of prospective prediction precision, prospective prediction sensitivity, F1 score, accuracy, and specificity at a 99% CI inspired by the previously reported method (*23*).

The anticipated clinical readout event proportion (φ) was set to 0.9 since, as of the cutoff date, an average of 75% of clinical trials on clinicaltrials.gov have reported clinical trial results in compliance with the FDAAA 801 and the Final Rule tracked by <u>fdaaa.trialstracker.net</u>. More than 90% of big-pharma-sponsored clinical trials report results (*40*).

The ratio of total observed clinical readout events and total expected clinical readout events (O/E) was set to 1 with a width of 0.15 to account for the 12% of clinical trials that are prematurely terminated without clinical trial results (*41*).

As such, the minimum sample size requirement was set to 131 predictions, and the minimal event size requirement was 118 readouts for a 99% CI. From February 11, 2020 to December 22, 2020, 265 prospective predictions were published to meet this minimal sample size requirement. By the time of the study cutoff date (March 1, 2022), 157 clinical readouts (5 phase 1 trials, 68 phase 2 trials, and 85 phase 3 trials) were available (Data S1). Thus, the minimum event size requirement has been met.

All other factors were held constant and performance measures were detected at a 90% CI for validation subgroups with smaller event sizes, using a minimum requirement of 52 predictions and a required event size of 47 readouts. Only simple means and standard deviations were calculated for validation subgroups with even smaller event sizes over corresponding proportions.

The direct translatability of prospective validations to clinical ABNN applications was evaluated using a two-sample Kolmogorov-Smirnov test, the standard approach for comparing validation clinical trials and initiated clinical trials since 2020. These results followed an identical distribution (Fig. 3A).

Statistical significance in real-world scenarios was ensured for actual clinical settings, using Agresti-Coull Intervals for the proportions and Wald Intervals for the differences (*42*). Two-sided p-values (*43*) were calculated for every proportion and difference and a 2% absolute margin was selected for non-inferiority and superiority comparisons, using a statistical significance threshold of 0.01. The SciPy and NumPy libraries in python were used for all function commands.

---

**Fig. S1.**
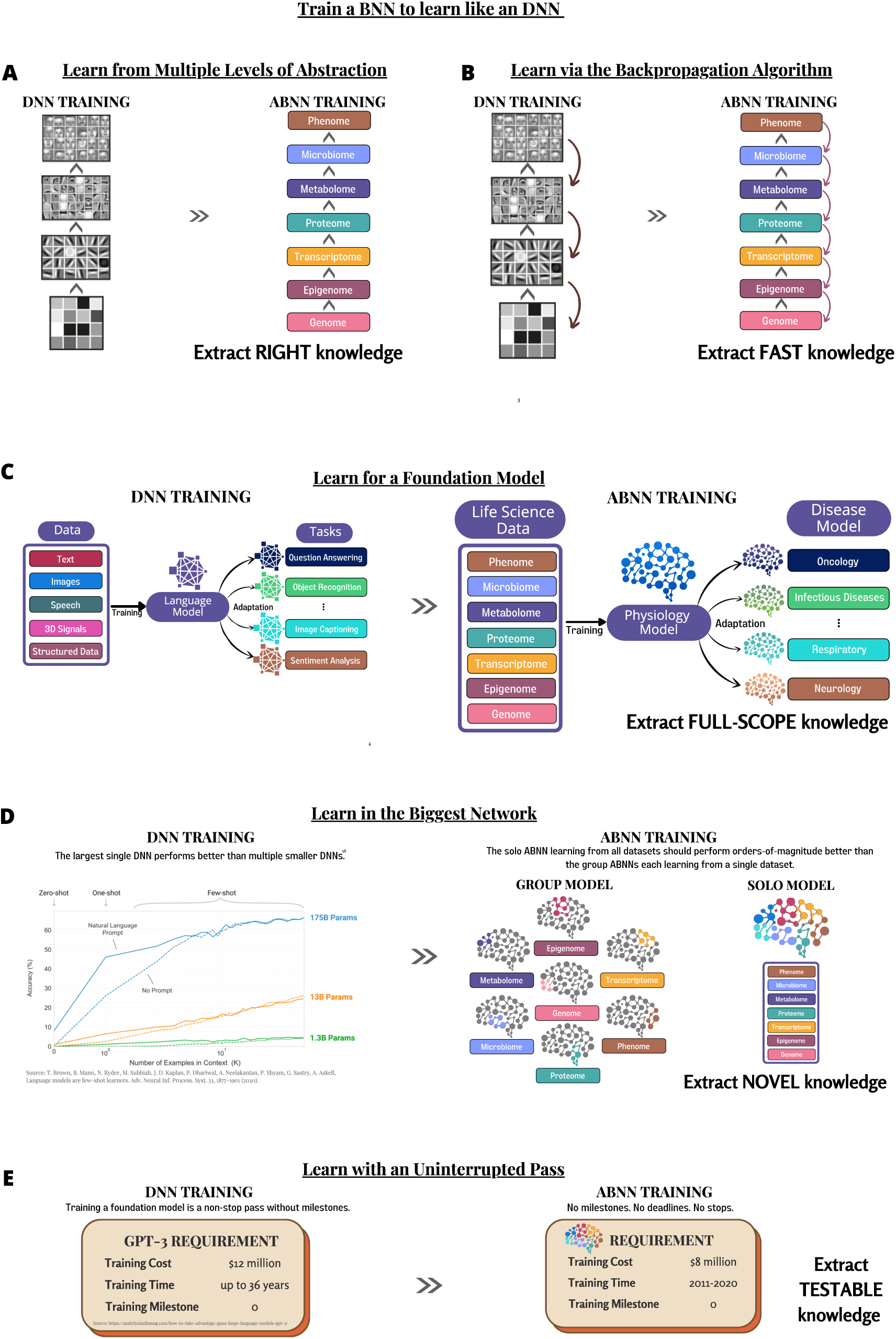
Train a BNN to learn like an DNN. (**A**) DNNs learn better representations of data with more levels of abstraction than with fewer levels and share a hierarchical architecture with BNNs. ABNNs extract right knowledge by integrating more multiomics data (e.g., genome, proteome, transcriptome, epigenome, metabolome and microbiome). (**B**) DNNs use backpropagation to improve learned representations of data by sequentially updating its internal parameters from the highest to the lowest levels of abstraction. The ubiquitous feedback connections in BNNs may be used to implement backpropagation-like learning rules. ABNNs extract fast knowledge by iteratively updating internal representations in a strict order from the most macroscopic level (phenotype), through the intermediate level (endophenotype), to the most microscopic level (genotype). (**C**) DNNs learn general-purpose representations of all-encompassing data to deliver optimal performance for a wide range of special-purpose tasks. Both BNNs and DNNs could learn to command general linguistic capabilities for multiple tasks. ABNNs extract full-scope knowledge by first learning a general-purpose model of human physiology from the all-encompassing data, prior to learning human diseases models from disease-specific data. (**D**) DNNs maximize the robustness of learned representations of data by increasing the sheer size of network parameters. ABNNs maximize extraction of novel knowledge if a single ABNN learns from the entirety of the all-encompassing data with neuronal nodes interconnected by always-on high-bandwidth biological synapses in a single brain, rather than multiple ABNNs learn from siloed sub-datasets with neuronal nodes interconnected by intermittently-on low-bandwidth human languages between several brains. (**E**) DNNs learn the best representations of data when provided with sufficient time and ample resources to allow for an uninterrupted pass of a full training set, prior to which no meaningful performance milestones should be defined. ABNNs extract testable knowledge if a milestone-free resources are provided for an uninterrupted learning pass, ensuring the ABNN won’t be evaluated prior to the completion of learning both human physiology and human diseases.

**Fig. S2.**
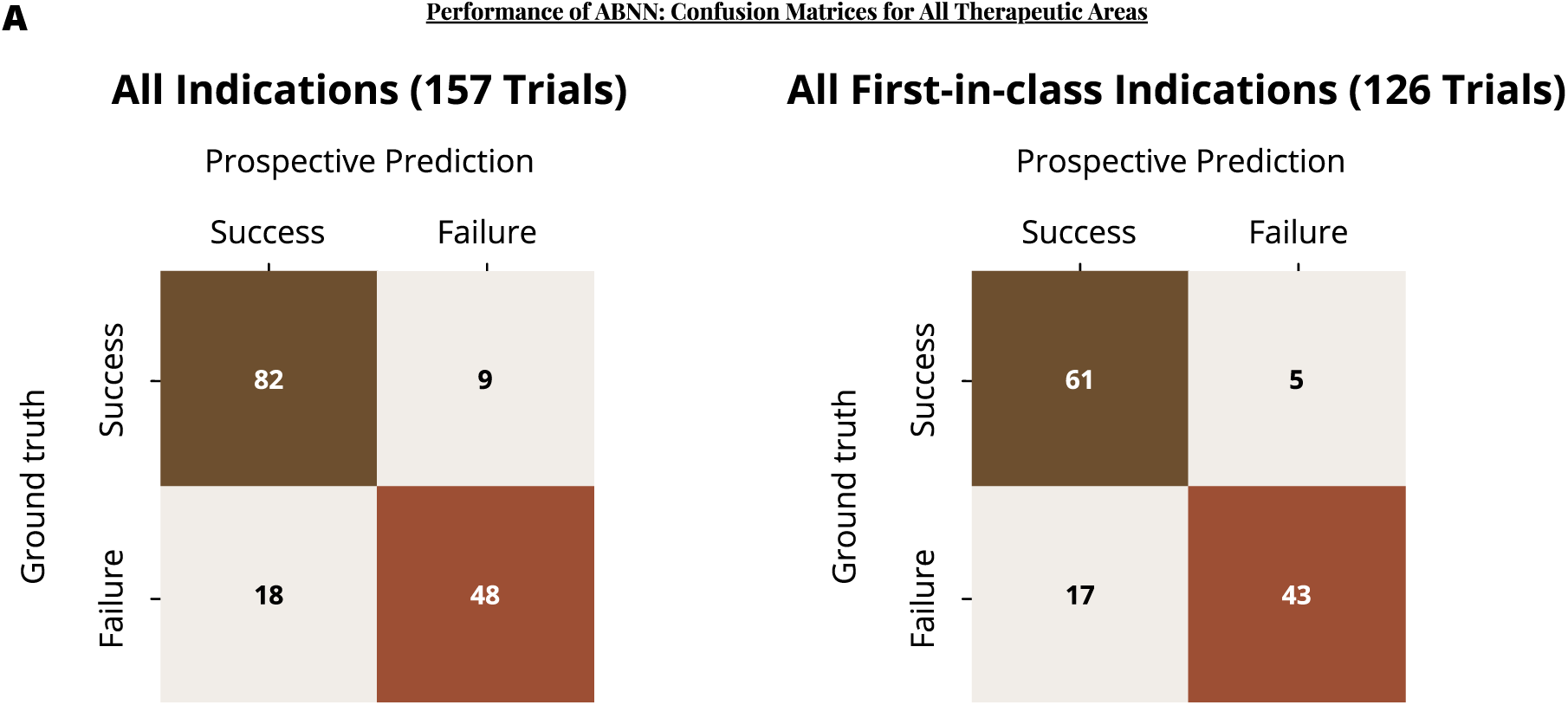
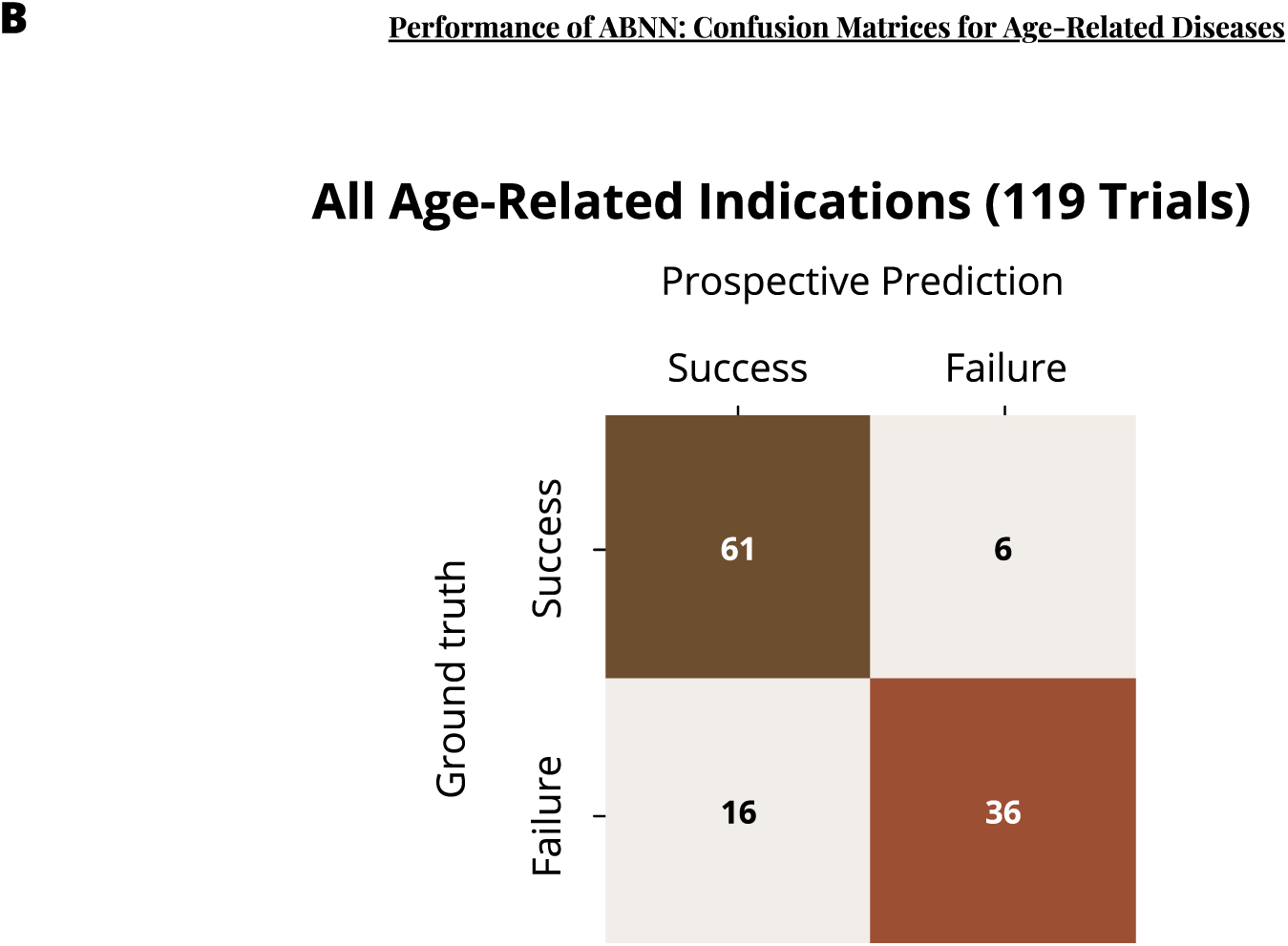
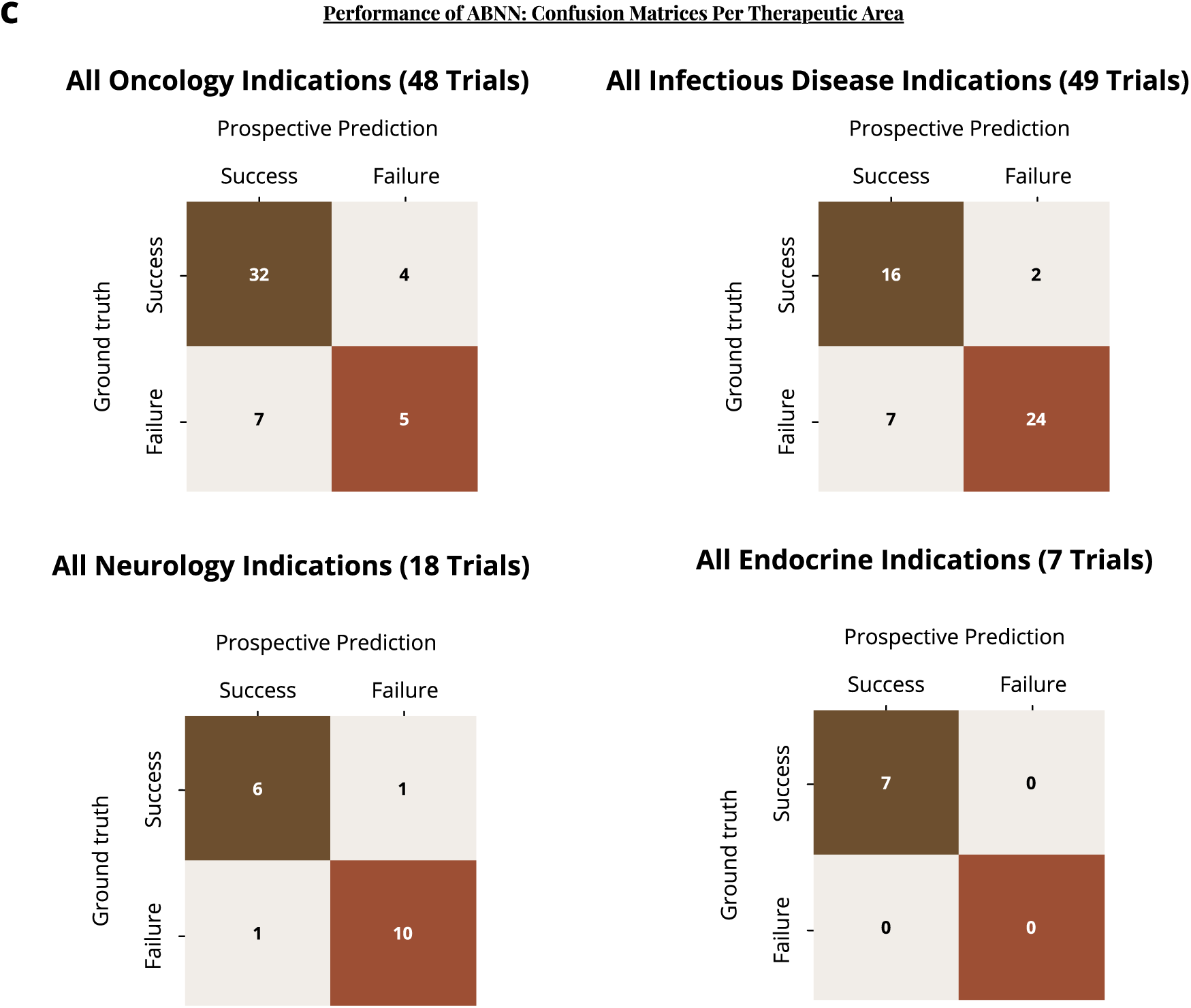
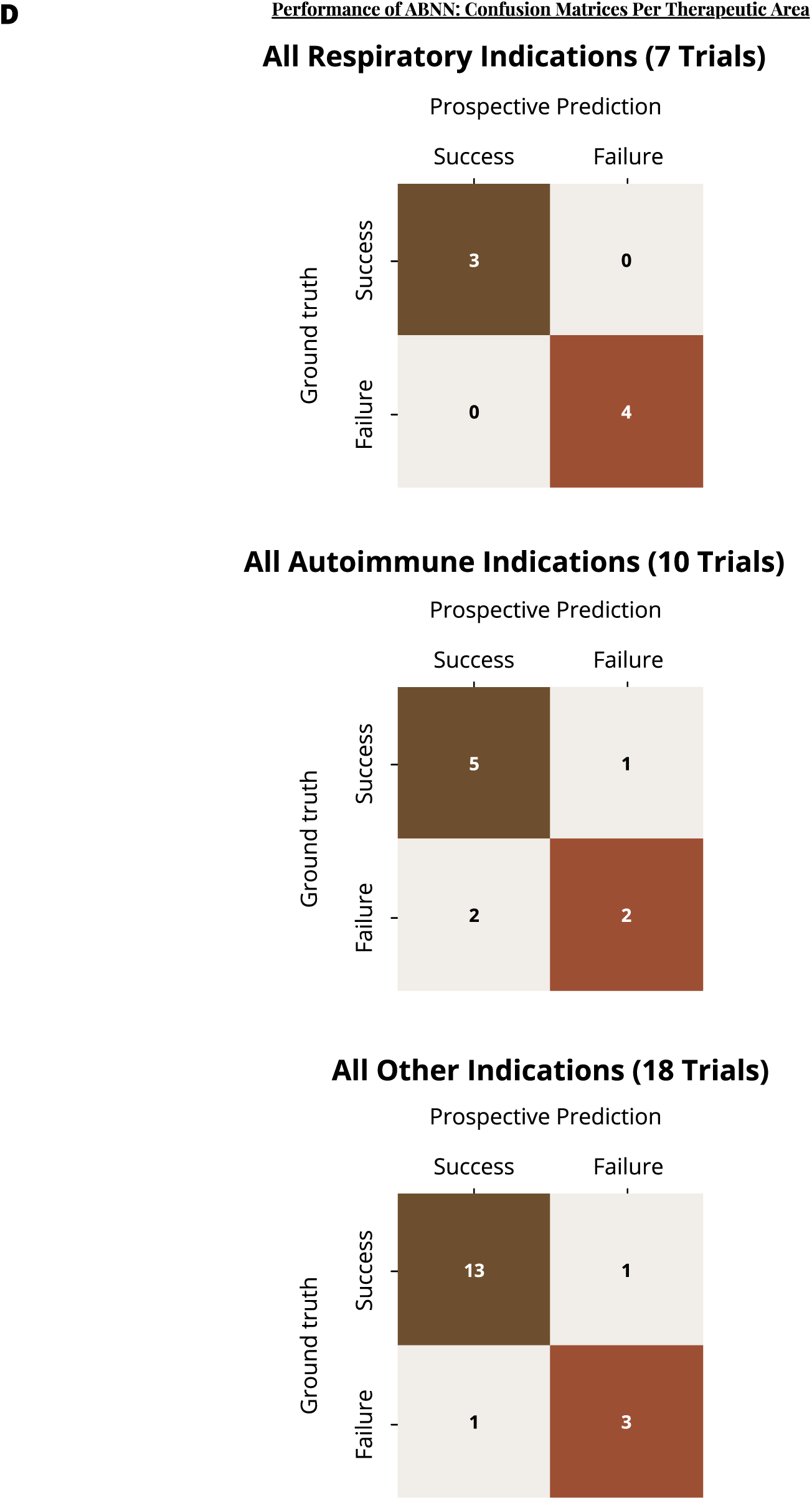
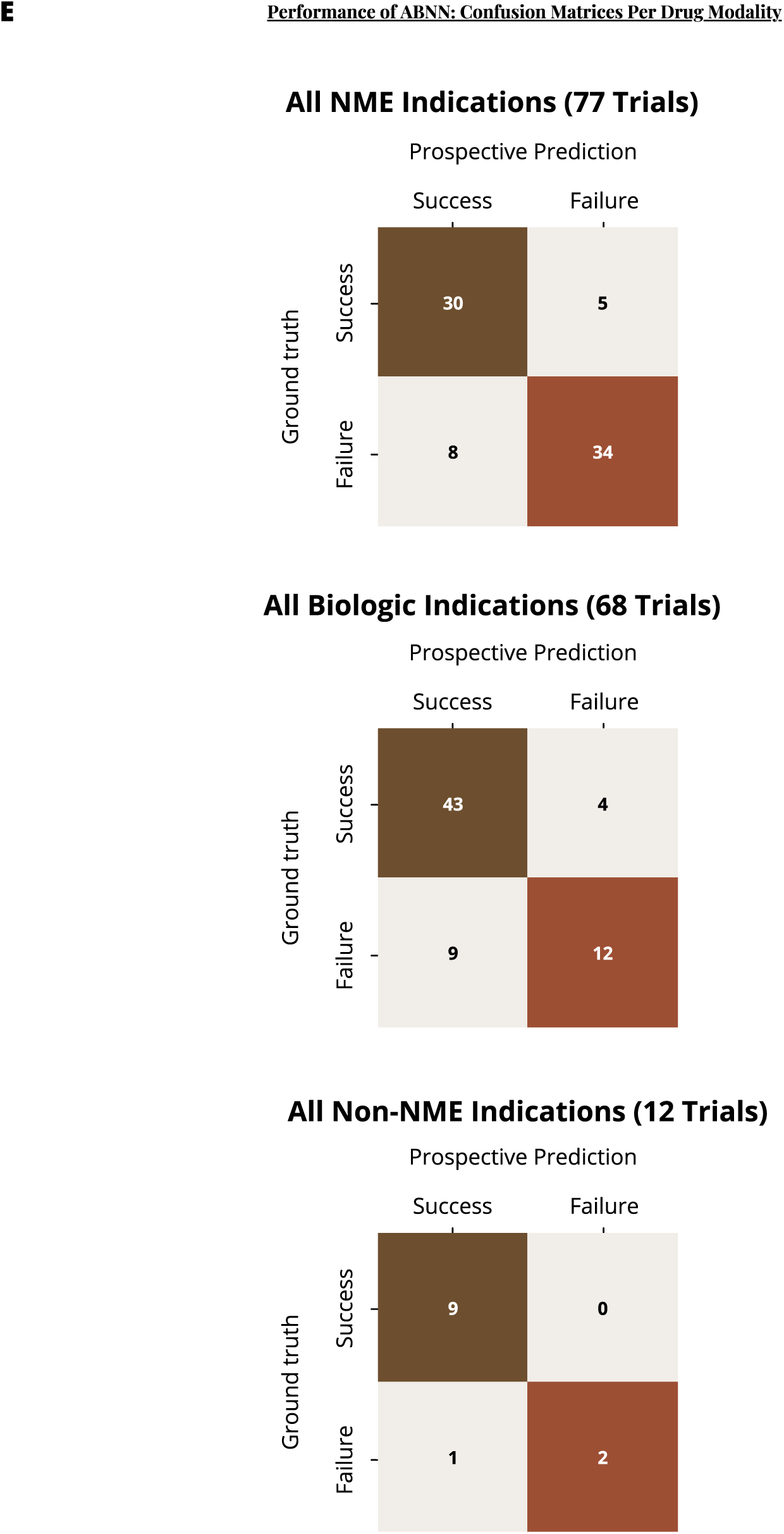
Confusion Matrices. (**A**) ABNN performance as recorded by confusion matrices for all therapeutic areas. (**B**) ABNN performance as recorded by confusion matrices for all age-related diseases. (**C**) ABNN performance as recorded by confusion matrices for respiratory, autoimmune, and other indications. (**D**) ABNN performance as recorded by confusion matrices for oncology, infectious diseases, neurology, and endocrine indications. (**E**) ABNN performance as recorded by confusion matrices per drug modality.

**Fig. S3.**
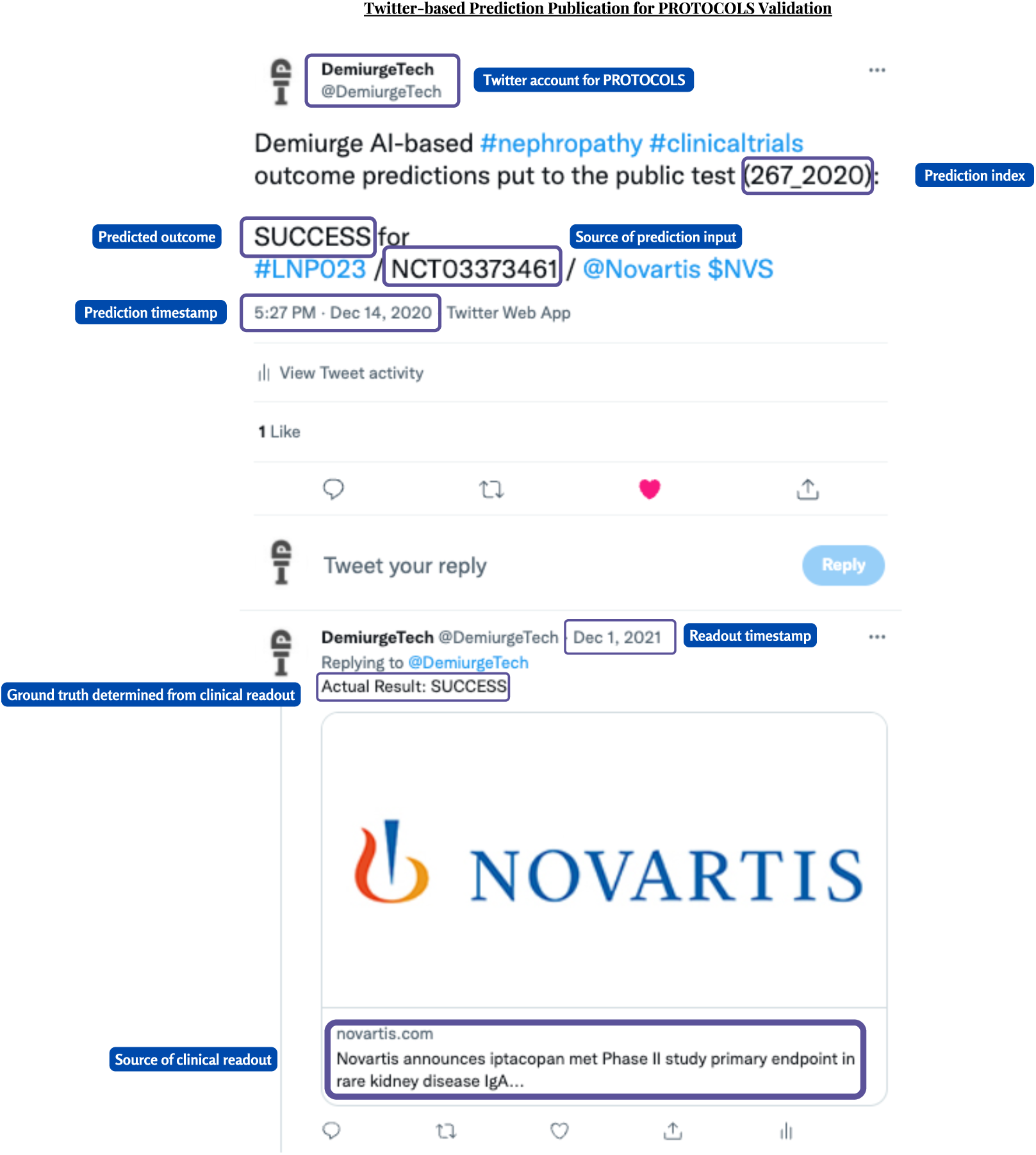
Twitter-based prediction publication for PROTOCOLS validation test. Each of the 265 predictions was published as a timestamped and indexed tweet @DemiurgeTech (see the annotated example above) to establish a publicly accessible track record for prospective prediction. The figure is a screenshot of the original tweets that can be accessed by visiting https://twitter.com/DemiurgeTech/status/1338536074654740481.

**Table S1.**
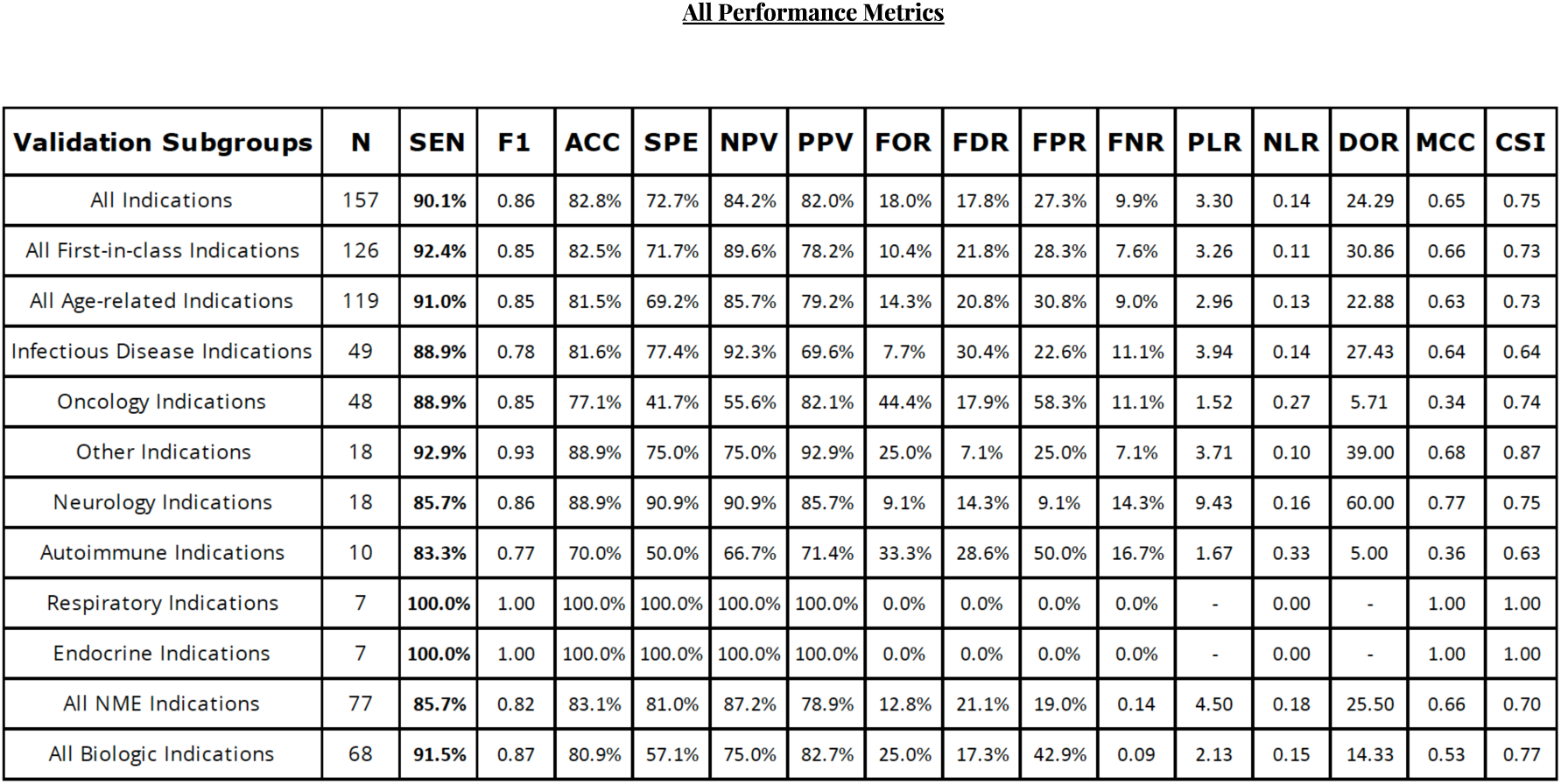
All performance metrics. SEN: Sensitivity; F1: F1-score; ACC: Accuracy; SPE: Specificity; NPV: Negative Predictive Value. PPV: Positive Predictive Value (Precision); FOR: False Omission Rate; FDR: False Discovery Rate; FPR: False Positive Rate; FNR: False Negative Rate; PLR: Positive Likelihood Ratio; NLR: Negative Likelihood Ratio; DOR: Diagnostic Odds Ratio; MCC: Matthews Correlation Coefficient; CSI: Critical Success Index; NME: New Molecular Entity (small molecule drugs).

**Data S1. (separate file)**

Detailed summary information of the PROTOCOLS validation data is available to download as separate Excel spreadsheets.

